# Advancing diagnosis of bipolar disorder using brain morphometric similarity networks in a graph AI framework

**DOI:** 10.64898/2026.05.12.26350596

**Authors:** Inês W. Sampaio, Giuseppe Poli, Alessandro Pigoni, Marcella Bellani, Francesco Benedetti, Igor Nenadić, Mary L. Philips, Fabrizio Piras, Jair C. Soares, Yvan Torrente, Lakshmi N Yatham, Anna Maria Bianchi, Eleonora Maggioni, Paolo Brambilla

**Affiliations:** Department of Neurosciences and Mental Health, Fondazione IRCCS Ca’ Granda Ospedale Maggiore Policlinico, Milan, Italy; Department of Electronics, Information and Bioengineering, Politecnico di Milano, Milan, Italy; Department of Neurosciences, Biomedicine and Movement Sciences, Section of Psychiatry, University of Verona, Verona, Italy; Division of Neuroscience, Unit of Psychiatry and Clinical Psychobiology, IRCCS Ospedale San Raffaele, Milan, Italy; Cognitive Neuropsychiatry Lab, Department of Psychiatry and Psychotherapy, Philipps-University Marburg, Marburg, Germany; Department of Psychiatry, University of Pittsburgh School of Medicine, Pittsburgh, PA, USA; Fondazione IRCCS Santa Lucia, Roma, Italy; Center of Excellence on Mood Disorders, Faillace Department of Psychiatry and Behavioral Sciences, McGovern Medical School, University of Texas Health Science Center at Houston, Houston, TX, USA; Department of Psychiatry, University of British Columbia, Vancouver, BC, Canada; Department of Pathophysiology and Transplantation, University of Milan, Milan, Italy

**Keywords:** Brain Similarity Networks, Morphometric Similarity, MIND networks, Brain Graphs, Neuropsychiatry, Bipolar Disorder, MRI, Deep Learning, Graph Neural Networks, GCN

## Abstract

Brain similarity networks (BSNs), extracted from structural magnetic resonance imaging, provide a validated framework for studying brain network organization and encode neurodevelopmental information relevant for psychiatric disorders. Recently, a neurodevelopmental hypothesis has been proposed for bipolar disorder (BD), where evidence demonstrates neuroprogression phenotypes differing from controls. BSNs offer a promising framework for investigating BD’s neural correlates but remain largely underexplored. Parallelly, graph neural networks (GNNs) have emerged as suitable deep learning models for exploiting network-level information. This study aimed to investigate BSNs for discriminating subjects with BD from controls within a GNN framework using the multi-site StratiBip network, composed of 605 controls and 501 subjects with BD. Leveraging advanced analysis tools, we developed a multi-site classification framework including: i) the state-of-the-art MIND algorithm for computing morphometric similarity (MS) networks based on gray matter volumes (GMV), ii) MS integration with age, sex, and GMV, iii) a leave-one-site-out cross-validation for multi-site model generalizability evaluation. The best model achieved a mean multi-site accuracy of 68%. Explainability analyses revealed meaningful MS patterns in the basal ganglia, frontal and temporal lobes, and a particularly relevant integration with age. This study provides interpretable insights into the role of MS in BD and unveils evidence supporting ageing-related processes as a significant component of BD pathophysiology.

## INTRODUCTION

Brain structural similarity networks have emerged as relevant alternatives to structural connectomes for studying the network organization of the brain (*1*). Morphometric similarity among brain regions, derived from structural magnetic resonance imaging (sMRI) data, offers a low-cost proxy for brain inter-regional microscale similarity and conveys robust information on typical and atypical regional neurodevelopment (*2, 3*).

This emerging approach is based on the assumption that common experience-related brain plasticity, such as that reflected in cortical differentiation, myelination, or lamination patterns, is encoded at the macroscale (*4*). Morphometric similarity thus unveils a representation of the brain *architectome* (i.e., architecture of the connectome), which acts as a proxy for the structural connectome. For this assumption, the morphometric inverse divergence (MIND) algorithm (*5*) stands as the most robust method to construct such networks, with evidence that MIND networks can be well interpreted as brain architectomes (*1*). Recent evidence shows that morphometric similarity has high sensitivity to age-related changes in brain organization, which has been reflected in their accurate prediction of an individual’s age from a single MRI scan, outperforming age prediction by DTI in the same sample (*5*). A link between brain morphometric similarity and brain functional networks has also been demonstrated, with inter-regional similarity in cortical thickness efficiently predicting functional connectivity in humans (*6*).

These unique properties have brought a growing interest in investigating brain morphometric similarity networks in complex psychiatric disorders (*7, 8*). This interest is largely motivated by the limited translational advances achieved with conventional neuroimaging approaches and by the unique perspective offered by brain similarity networks for investigating disorders for which a brain dysconnectivity hypothesis has been proposed, such as the two major psychoses, bipolar disorder (BD) and schizophrenia (*9, 10*). BD has proven particularly challenging to predict using structural neuroimaging. Although numerous studies have been conducted in the last decades, findings remain fragmented, leaving an urgent need to identify reliable biomarkers and develop objective and biologically driven diagnostic frameworks for BD. Structural brain alterations have been identified, but their predictive power is limited (*11, 12*), and group-level findings often fail to replicate well at the individual level (*13, 14*).

In this context, brain morphometric similarity is promising, due to its cost-effectiveness and neurodevelopmental focus. To date, in psychiatry, morphometric similarity has been explored in schizophrenia and autism spectrum disorders, both of which are hypothesized to involve disrupted neurodevelopment processes (*15, 16*), as well as in paediatric BD (*17*). However, the utility of morphometric similarity for BD remains largely underexplored. A recent hypothesis points towards a neurodevelopment model of BD (*18*), and accumulating evidence suggests that BD might be a disorder associated with accelerated brain ageing, stemming from replicable findings of different neuroprogression in subjects with BD when compared to healthy controls (HC). Thus, morphometric similarity profiles provide a sensitive framework for capturing structural and network-based disruptions underlying this complex disorder.

Parallelly, the search for brain network markers of BD can particularly benefit from the enormous advances in graph-based AI analytics. The introduction of graph neural networks (GNN) has provided powerful tools for studying network-structured data, offering a framework to exploit the complex structure of brain morphometric similarity networks. This is particularly relevant for BD, where subtle and distributed network alterations may not be detectable using conventional univariate or graph-theory-based approaches.

In this light, this study aimed to investigate the effectiveness of a GNN analysis framework based on brain morphometric similarity networks in discriminating subjects with BD from HCs. We leveraged the improved MIND algorithm to extract robust individual-level morphometric similarity networks based on gray matter volume (GMV) MRI maps. We entered the MIND networks and complementary information within a new, robust, and explainable GNN framework. We applied this framework to data from the multi-site StratiBip cohort, composed of more than 1000 subjects balanced between controls and subjects with BD, to advance diagnosis and inform on quantitative and generalizable brain network patterns of disease.

Our methodological contributions include the proposal of a robust multi-site GNN classification framework incorporating a leave-one-site-out cross-validation (LOSO-CV) scheme with an innovative, integrated multi-site harmonization strategy between training and independent test sets. In addition, we conducted a series of sensitivity analyses to evaluate the integration of MIND similarity profiles with demographic variables (age and sex) and regional GMV metrics. Finally, we rigorously assessed the clinical validity of the resulting GNN model through a comprehensive set of explainability (xAI) analyses.

We hypothesized that MIND networks would encode stronger predictive information than metrics of brain regional morphometry alone, potentially capturing links between neurodevelopmental processes and neuroprogression across the lifespan that could play a relevant role for BD. Furthermore, we hypothesized that leveraging a state-of-the-art GNN framework in a multi-site research context would further enhance the prediction of BD by increasing diagnostic accuracy and generalizability.

## RESULTS

### Pipeline Overview

In Fig. 1, we present a general overview of the analysis pipeline employed in this study. We used structural magnetic resonance imaging (sMRI) data from 1106 subjects from the multi-site StratiBip dataset, composed of BD and HC, to compute subject-level brain morphometric similarity networks, based on the MIND algorithm (*19*), which hereinafter will be called MIND networks. Here, the MIND networks represent the pairwise GMV similarities between regions of interest (ROIs) from the neuromorphometrics brain atlas (N=122 ROIs) (*20*). Then, MIND networks were harmonized across sites, and those from a subset of subjects were kept to balance age distributions between HC and BD (details in Supplementary Materials). The selected MIND networks were subsequently fed to a GNN classification analysis for discriminating subjects affected with BD from HCs. In a first stage, a series of sensitivity analyses aimed at evaluating the influence of several graph and model design choices on model performance, reducing the hyperparameter search space and improving the final classification framework. These trials explored multiple combinations of: i) GNN and ML models with varying numbers of layers and hidden dimensions, ii) graph node features (MIND values, age, sex, ROI GMV), iii) graph densities, and iv) node normalization techniques. The best combination sets were passed to the last stage of the pipeline, focused on model validation, where we employed a leave-one-site-out cross-validation (LOSO-CV) framework to assess the model’s generalizability across independent acquisition sites. The best results were compared to traditional ML methods, and explainability methods were used to extract a clinical interpretation of the results. Our code is available on GitHub: https://github.com/GiuseppePoli/MBNGNN.git.

**Fig. 1.**
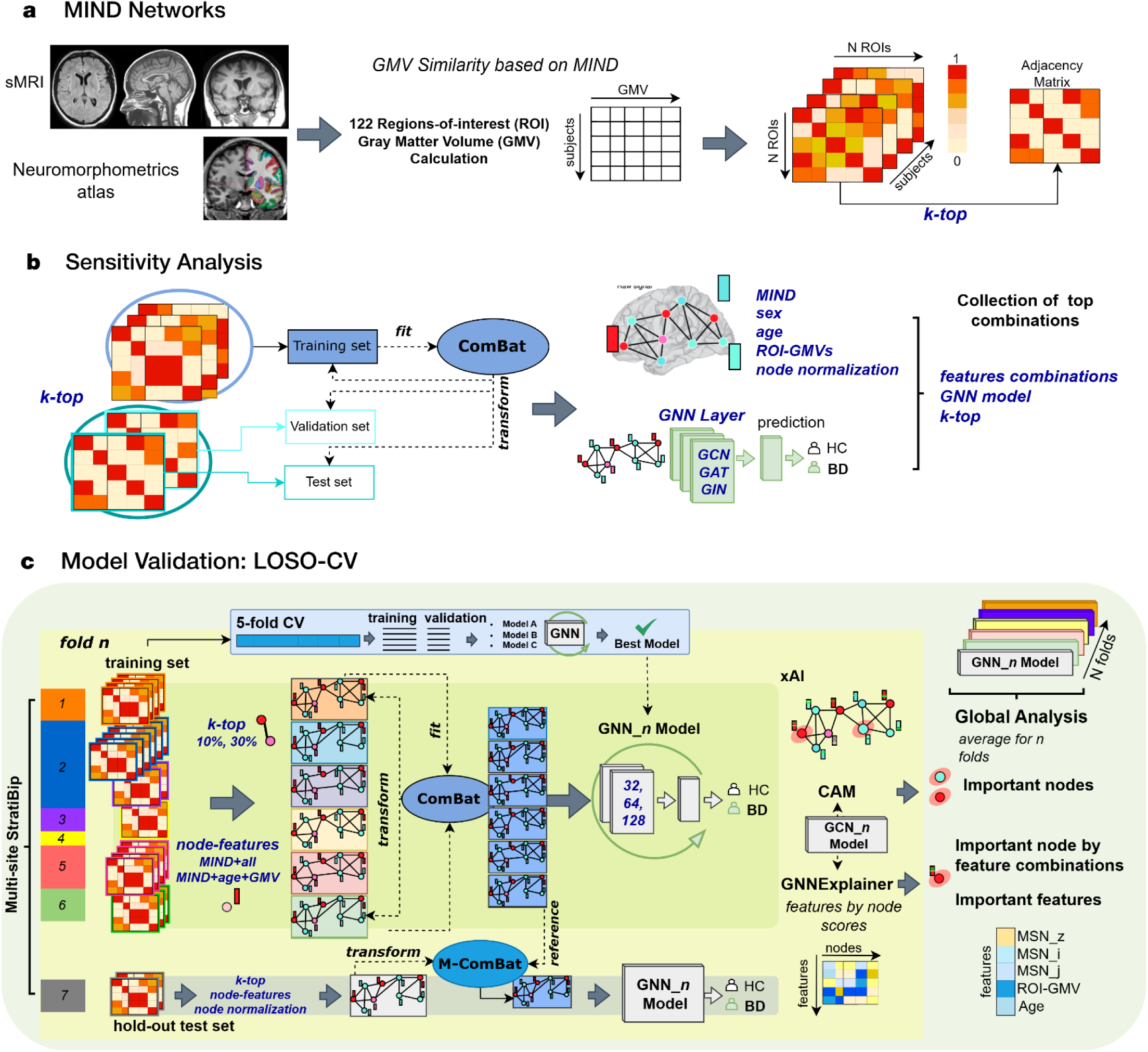
Analysis pipeline overview. Calculation of the MIND networks based on GMV from sMRI data. A set of sensitivity analyses was employed to study different graph designs and model configuration choices and their impact on the classification task with GNNs. Finally, a model validation stage based on a LOSO-CV was used to select the final graph design and best model configuration and assess the model’s generalizability across different sites. xAI techniques were employed at the fold level and then aggregated and summarized, where global node and feature importances were evaluated. Text in blue describes the set of hyperparameters that were tested in the sensitivity analysis and in the LOSO-CV.

### Sensitivity Analyses

The sensitivity analyses consisted of several experiments regarding the model and graph characteristics. After age balancing, the entire dataset was composed of 544 HC (median age = 33 years, 307 females, 237 males) and 453 BD (median age = 39 years, 265 females, 188 males). The dataset was partitioned into three distinct subsets: training (70%, 774 subjects), validation (15%, 166 subjects), and test (15%, 166 subjects), stratifying for site, i.e., the proportion of subjects for each site was kept in the three subsets. The ComBat model (*21*) was used to harmonize the MIND networks, respecting the independence between train, validation, and test sets, avoiding any data leakage (*22*). The harmonization effectiveness was visually assessed, as presented in Fig. S1. Different model configurations and graph design choices were compared in their performance on the validation set to select the best-performing configuration, which was then tested on the unseen test set to compute the final classification accuracy.

The first analyses included the comparison between GNN models and ML models, revealing the superior performance of the graph convolutional neural network (GCN) model (Table S1). Following, using the GCN model, we probed different node feature combinations including MIND values, age, sex and ROI GMV. MIND showed higher predictive power when compared to ROI GMV and demographic variables, and its combination with age and ROI GMV stood out as the best performing node feature set (Table S2). Next, we assessed the impact of graph density in model performances, testing sparser graphs with top 10% and 30% of edges, and denser graphs with top 70% and 90% of edges. We found that sparser graphs achieved satisfactory and comparable results with more denser graphs, with the advantage of reducing training time due to its lighter structure (Table S3). Finally, we performed hyperparameter tunning on the GCN, evaluating varying number of layers, hidden features and node normalization methods. The GCN with 2 layers achieved better average performance and had the least computational burden (Fig. S2), therefore it was used for the validation phase.

### Model Validation: Leave-One-Site-Out Cross-Validation

A GCN model with 2 layers and multiple hidden dimensions was evaluated under a LOSO-CV. Iteratively, data from one site was held out as a test set while all others were considered in a 5-fold CV framework as a training set. The 5-fold CV was used for hyperparameter tuning, where the GCN model was trained in the internal training set and evaluated in the internal validation set. The hyperparameter tuning was performed considering varying hidden dimensionalities of 32, 64, and 128 features. Additionally, in this stage, we evaluated the four best node normalization strategies: standardization (norm_std), detrending (norm_mean), dividing by the square root of standard deviation (norm_rsd), and scaling with p-rooth of the variance (norm_pr_v). The best model architecture was then retrained in the entire training set and tested in the hold-out data. All sites, except site 2, underwent this process, where their data was used once as a test set for model evaluation. Site 2 comprises a large portion of the StratiBip dataset; thus, we hypothesized that the training set’s dependency on this site was non-negligible. The age balancing procedure and multi-site data harmonization were applied within each fold of the LOSO iterations (*Fig. S3-S8*). Here, the graph structure and topology were kept fixed across evaluations, using the best node feature combination, MIND+age+ROI GMV, and MIND+all, with graph density levels of 10% and 30%. Thus, the GNN model configurations were evaluated for these four fixed conditions, tested across the LOSO-CV folds.

#### GCN Classification

Fig. 2 shows the accuracies obtained for each hold-out site in the LOSO-CV for the four models. The configuration combining a 10% graph density with MIND+age+ROI GMV as node features achieved the highest mean accuracy of 68.07%. Further performance metrics are reported in Table 1. This GCN model outperformed other baseline ML models, showing higher stability across all sites (Table S4). Nevertheless, a Friedman test that compared the four models’ performance across sites did not reveal any significant differences, although the test had very low power (n=6) and high variance, which introduced substantial limitations in the statistical test. The performance variability across sites underscores the challenges of multi-site generalization and demonstrates the importance of evaluating it. The LOSO-CV framework enabled us to identify a pipeline configuration that consistently generalized across sites. This is particularly evident for site 5, which produced poor performances for 3 out of 4 configurations. In the 3 failed configurations, the model classified all subjects as healthy, with an accuracy of 33.50% that exactly represents the percentage of HC in this site. We assessed the impact of not harmonizing the test set, showing comparable results for all sites and models, except for one site. The results, described in the Supplementary materials, demonstrate that, when demographic distributions and sample sizes are sufficiently aligned between the sites in the training and test sets, the model can generalize with different graph configurations. Differently, in the case of site 7, the model achieved consistently lower performances, ranging from 45.88% to 56.47% of accuracy. We hypothesize the age difference of this cohort as one explainable variable for the poor performance. The age distribution of site 7 differs substantially from that of other sites, with an average age of 22.72 years, while the remaining training set in this fold had an average age equal to 39.59 years. This age mismatch induced the model to be tested in an out-of-distribution scenario, where the model’s generalization achieved its critical limit. On the other hand, performances like the one obtained for site 3, ranging from 71% to 81.6% of accuracy, were potentially inflated, as this was the site with the smallest cohort in the dataset. Sites 1 and 6 provided more stable performances across trials, and in both cases, the configurations without sex provided the best results.

**Fig. 2.**
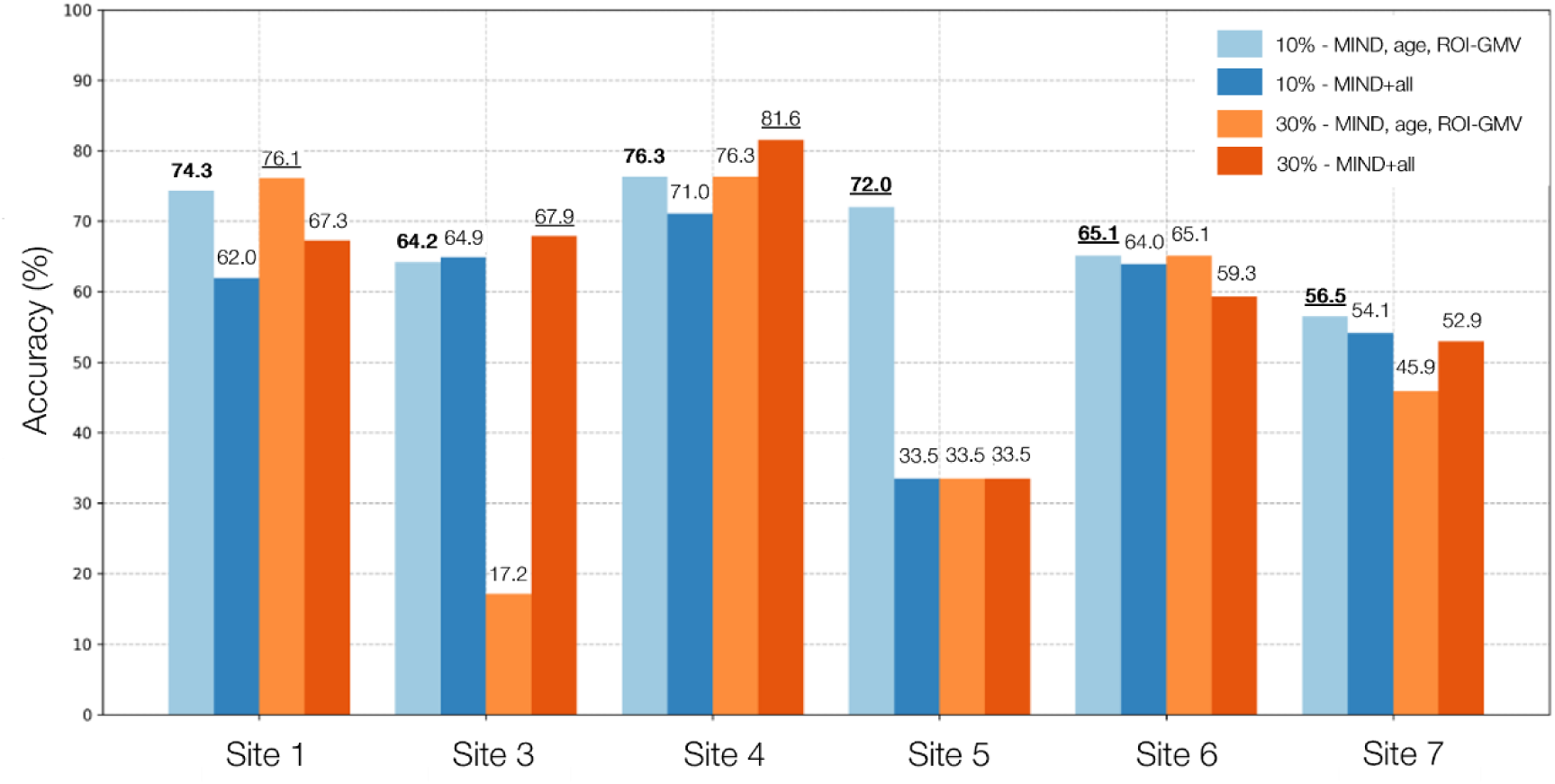
LOSO-CV performances across the four model trials. The accuracy results for the best overall model are represented in bold, while the best result for each site is represented with underlying.

**Table 1.** Detailed results from the best LOSO-CV trial (light blue in Fig.5), with a GCN model using graphs with 10% density and MIND+age+ROI-GMV nodes features.

| LOSO-CV | Accuracy(%) | Precision | Recall | TN | FP | FN | TP |
| --- | --- | --- | --- | --- | --- | --- | --- |
| Site 1 | 74.34 | 38.46 | 75.00 | 69 | 24 | 5 | 15 |
| Site 3 | 64.18 | 28.07 | 69.57 | 70 | 41 | 7 | 16 |
| Site 4 | 76.32 | 63.64 | 58.33 | 22 | 4 | 5 | 7 |
| Site 5 | 72.00 | 77.78 | 73.68 | 46 | 21 | 35 | 98 |
| Site 6 | 65.12 | 71.70 | 65.52 | 18 | 10 | 20 | 38 |
| Site 7 | 56.47 | 59.34 | 49.09 | 21 | 9 | 28 | 27 |
| Total Average | 68.07 | 56.50 | 65.20 | 41 | 18.2 | 16.7 | 33.5 |

### GNN Model Explainability

An explainability analysis was performed for the best model in the LOSO-CV, i.e., the GCN model with 10% density graphs and MIND+age+ROI GMV as node features. The LOSO-CV fold results were aggregated and averaged to compute the overall feature importance. Two techniques were used, the class activation map (CAM) to evaluate the contribution of the graph nodes (brain ROIs), and the GNNExplainer to evaluate the contribution of each nodal feature within each node, increasing the granularity of our analysis. A comparison with baseline statistical testing is presented in Tables S6-S7.

**CAM.** Fig. 3a) shows the average graph node importance. The right and left cerebellum exterior obtained the highest rankings in the brain network. Following, the nodes with the highest importance were in the bilateral temporal and frontal lobes, including middle temporal gyrus, middle frontal gyrus, and inferior temporal gyrus.

**Fig. 3.**
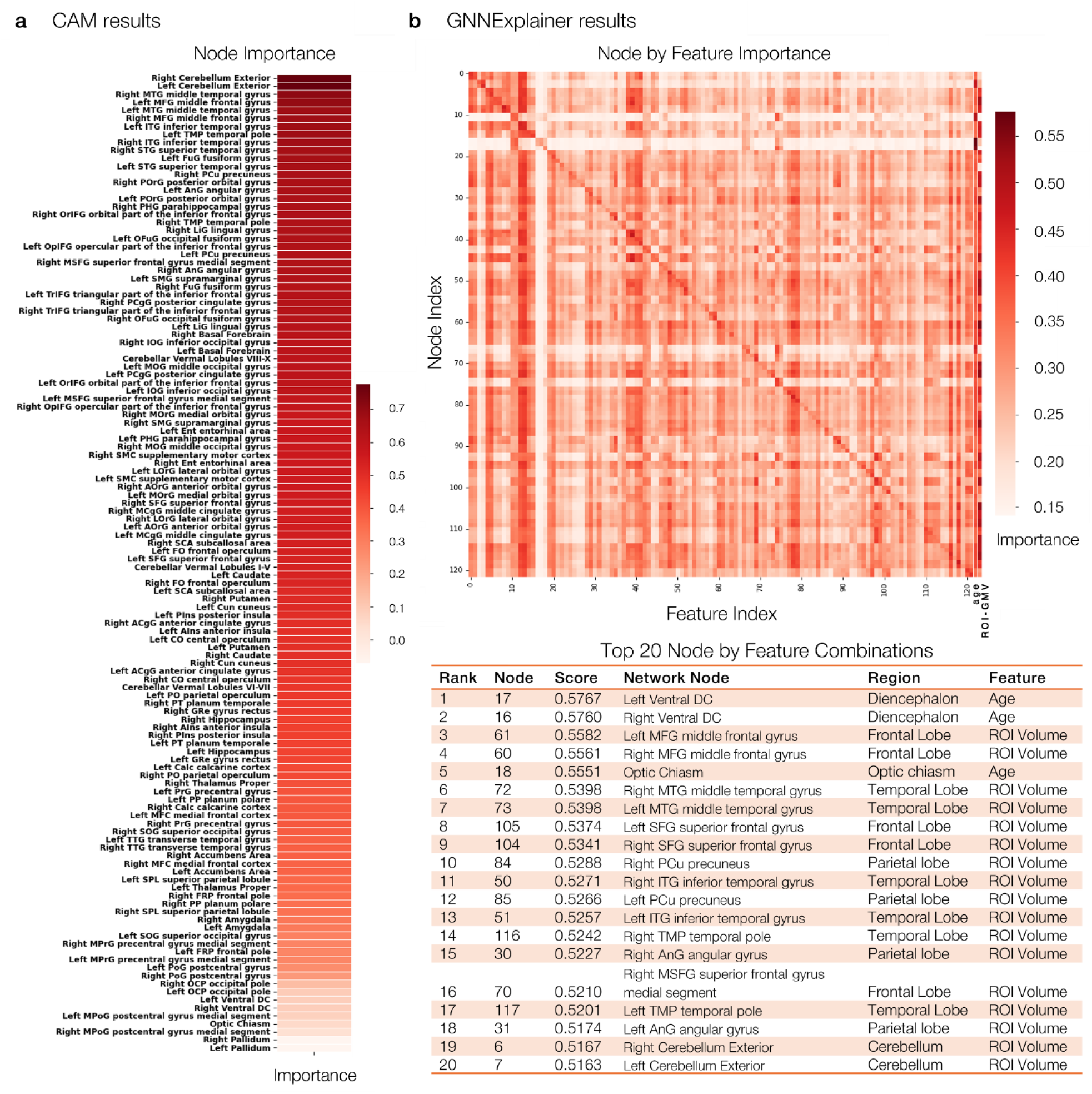
Average xAI results from the best LOSO-CV model. a) CAM results are plotted in a sorted heatmap representing the average graph node importance for discriminating HCs from subjects with BD. b) The GNNExplainer results are shown in two figures. On top, the heatmap representing the importance of all the graph nodes (row) by feature (column) combinations. The rows graph nodes IDs (122 ROIs) and columns feature IDs (total 124: MIND for the 122 ROIs, followed by age and ROI GMV) corresponding labels are shown in *Table S5*. At the bottom, top 20 graph node – features combinations.

**GNNExplainer.** Fig. 3b) shows the node-feature combination importance heatmap. Features that were consistently important for all nodes included age, ROI GMV, and MIND values of bilateral putamen and caudate, right temporal pole, bilateral frontal operculum, and bilateral opercular part of the inferior frontal gyrus. The pairs of node-feature combinations with the highest importance included the left and right ventral diencephalon as nodes, and age as feature (Table in Fig.3b). This suggests a strong relevance of age-related variations in these regions in patients with BD. Following, the top-ranking node-feature combinations were given by ROI GMV as feature in nodes belonging to the parietal, frontal, and temporal lobes. The cerebellum exterior nodes, which were found to be the most important in the CAM analysis, ranked 18th and 19th in combination with ROI GMV as feature. To obtain a global feature importance, we extracted the features having top-ranked mean importance scores over the graph nodes (Fig.4). The ROI GMV ranked first (mean score of 0.41), whereas age ranked third, supporting the relevance of age integration with brain morphometric similarity found in the sensitivity analysis. Particular importance for BD classification was found for MIND profiles of the basal ganglia, especially the putamen and the caudate, and of frontal regions —especially with the rest of the brain, represented by the graph nodes.

**Fig. 4.**
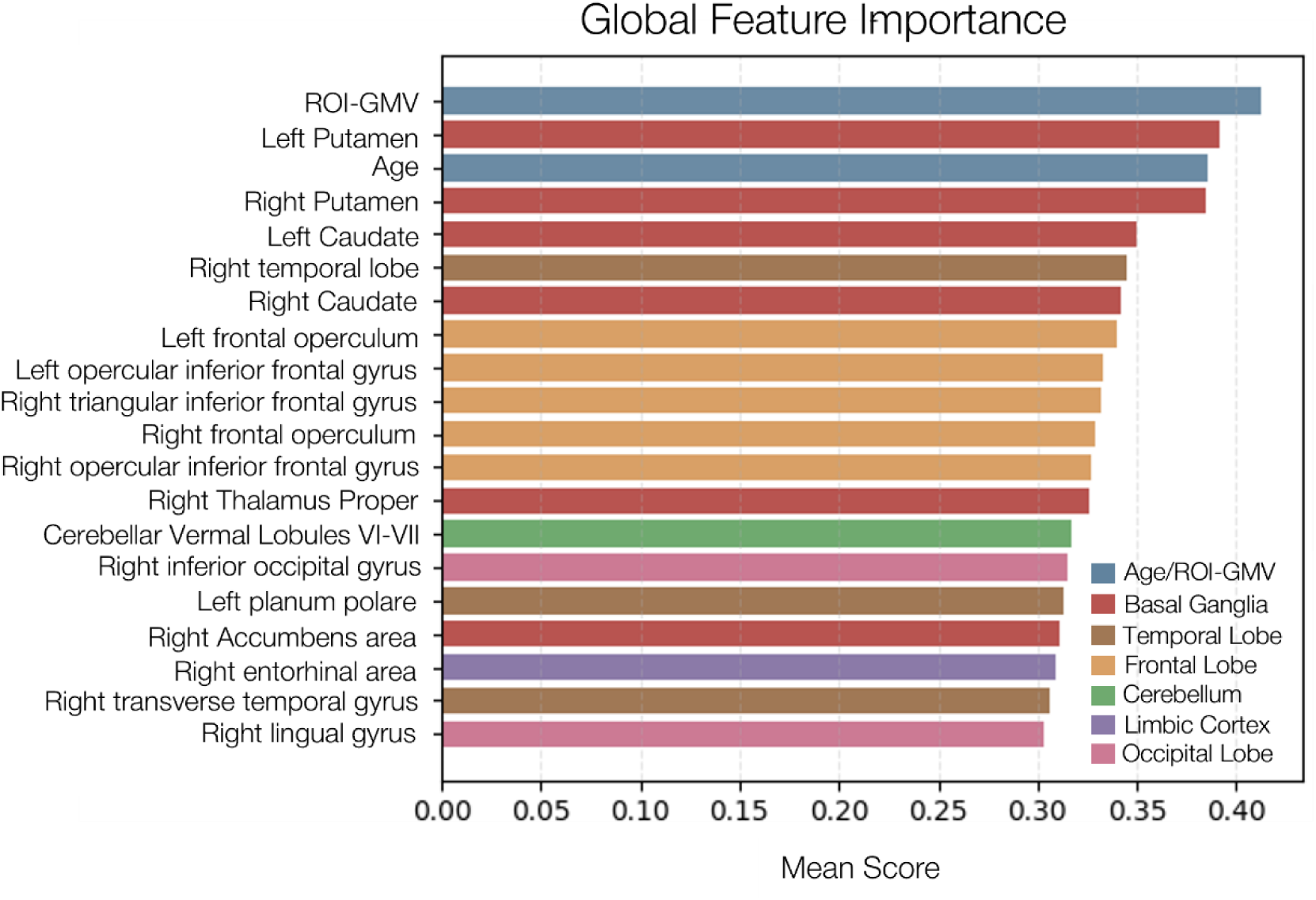
Average node feature scores from GNNExplainer. The plot shows the top 20^th^ node features in terms of global importance for all graph nodes in the GCN model.

## Discussion

In this study, we implemented a robust, generalizable, and explainable GNN framework based on brain GMV similarity networks constructed using the MIND algorithm. For the first time, we tested this framework to assess the potential of morphometric similarity for discriminating individuals with BD from controls, using the multi-site sMRI dataset from the StratiBip cohort, composed of over 1000 subjects.

Our pipeline included an embedded multi-site harmonization step fully integrated into the CV frameworks, designed to address the heterogeneity of our multi-site sMRI dataset. We performed a comprehensive assessment of GNN model architectures and brain graph designs using a set of sensitivity analyses, culminating in a model validation stage based on a LOSO-CV framework. This enabled us to evaluate site-specific model performance and its variance between sites and highlight the challenges of multi-site generalizability and clinical transferability. Finally, we applied GNN explainability techniques to the resulting model’s decision process to empower an exhaustive investigation of the model’s scientific validity and clinical relevance. The application of this robust framework to the multi-site StratiBip cohort allowed us to advance the automatic diagnosis of BD compared to the most recent literature (*12*), achieving an average multi-site accuracy of 68%, and inform on candidate brain network markers for this complex and heterogeneous disorder.

The results suggest that MIND networks have greater predictive power than traditional brain structural features, such as regional GMV alone, and highlight the potential of GCNs to effectively leverage MIND networks for advancing machine-learning (ML)-based diagnosis. Integrating MIND similarity profiles with GMV and age, but not sex, at the node-feature level improved the models’ discriminative capabilities. It is important to note that GMV was corrected for total intracranial volumes (TIV), partly correcting for sex effects, and that the dataset was balanced for age. These findings suggest a meaningful diagnostic value of GMV and age when integrated with MIND profiles, but not when included alone, within the brain similarity graphs. This unveils a new discriminative pattern for BD prediction and aligns with evidence for altered age-related brain processes in BD (*18, 23*).

From a methodological perspective, we improved the exploitation of brain morphometric similarity by leveraging the innovative MIND networks within a generalizable and explainable GNN framework. For the first time, this framework was applied to discriminate BD from HC using the large multi-site StratiBip dataset. Using a tuned GCN model built from MIND networks, we outperformed previous studies that used ML models with regional morphometric features as predictors (*12, 24*). While the observed classification performance is moderate and does not reach clinical relevance thresholds, it aligns with findings for BD when large-scale multi-site datasets are employed (*25*). For example, a large-scale ENIGMA study examined cortical thickness, surface area, and GMV features in controls and subjects with BD from over 13 sites, achieving a mean classification accuracy of 58.67% in a LOSO-CV framework (*12*). The low predictive power for BD is also supported by sMRI evidence of less prominent differences between BD and HC, when compared to schizophrenia (*26, 27*), and partly overlapping patterns between BD and HC (*14, 26*). Of note, the moderate accuracy obtained in our study may also be due to our decision to include BD individuals with heterogeneous clinical profiles in a single patient cohort. We deliberately did not stratify by clinical dimensions in order to assess the predictive potential of MIND networks for BD as a whole, demonstrating the existence of neural bases of BD with relevant clinical translational potential.

Using GNN explainability techniques, we identified the most important predictors for our model. From the MIND networks, we found that both nodes and node features in the cerebellum, basal ganglia, temporal lobe, and frontal lobe were the most relevant for discriminating subjects with BD from HC. Specifically, with CAM, we found that the bilateral exterior cerebellum nodes were the most important. Previous studies found the cerebellum to be involved in emotional regulation and social cognition (*28*), being significantly impaired in BD (*29*), and prior studies have identified reductions in GMV in this region in BD (*30*), also compared to schizophrenia (*31*). Notably, the cerebellum is considered an important region for suicidal behaviours, more prevalent in BD than in HC, and has been identified as predictive of suicide attempts or high risk for suicide in BD cohorts (*32, 33*).

A more granular analysis with GNNExplainer allowed us to inspect relevant node-feature combinations. The highest importance score involved the bilateral ventral diencephalon in relation to age. In the neuromorphometric brain atlas, this region includes the hypothalamus, mammillary body, subthalamic nuclei, substantia nigra, red nucleus, lateral and medial geniculate nucleus, structures that contribute to the limbic, basal ganglia, and motor control systems. Previous normative studies found GMV of the ventral diencephalon to be quadratically related to age (*34*), hence our findings suggest that BD pathophysiology involves altered brain ageing processes for this region. Additionally, as demonstrated in the sensitivity analysis, combining age with MIND values and GMV achieved the best classification performance. This indicates that, besides the effect of age on GMVs throughout the normative lifespan, there is a non-normative interaction between age, GMV, and MIND similarities in BD. In the same line, Nunes et al. 2020 report that age, and not sex, was associated with being classified as BD by their ML model.

In this context, several studies have reported altered age effects in BD, like accelerated aging (*23*), which is reflected in different dimensions, such as progression of cognitive decline and executive functions, and higher brain-age estimations. However, findings are fragmented, as some studies reported a worse cognitive decline in BD than in HC, while others reported equal progression of age-related cognitive decline in both groups (*35, 36*). One study reported advanced brain aging in BD (*37*), while another found no differences to HCs (*38*).

Our study supports the hypothesis that age could have a different impact on some brain structures in BD compared to HC. In our age-balanced dataset, we found that age combined with other neuroanatomical features played a significant role in predicting BD. Indeed, age ranked third in terms of node feature importance, after regional GMV and left putamen MIND profile. Although the involvement of the putamen in BD pathophysiology is recognized (*39*), the basal ganglia are known to be greatly impacted by the cumulative use of antipsychotics (*40*), and we did not disentangle the effects of medication from those of the disease.

Furthermore, the most important node-by-feature combinations consistently included MIND values of the frontal and temporal gyri. Similarly, the top-20 global features were mainly represented by the MIND similarity profiles of regions in temporal and frontal lobe and in the basal ganglia.

These results align with our baseline analysis employing statistical testing to the MIND similarity profiles (Fig. S10 Table S6-S7), and with literature on GMV alterations in BD. This literature has reported lower thalamic volumes, as well as cortical alterations mainly in frontal and temporal areas (*11, 27, 41*). Several studies have also reported greater GMV in basal ganglia structures, such as the putamen and caudate, in subjects with BD and schizophrenia (*42–44*). The inferior frontal gyrus has frequently been reported as altered in BD, both functionally (*45*) and structurally (*46*). Nevertheless, our results cannot be interpreted as independent regional evidence, but as part of a network-based pattern that explains how the model discriminated HC from subjects with BD.

To the best of our knowledge, only one study has investigated morphometric similarity networks in BD in a much smaller paediatric sample (*17*). Consistent with our and previous findings, they found differences in morphometric similarity networks mainly in frontal and temporal cortices. Specifically, they observed lower similarity in the bilateral dorsomedial prefrontal cortex, left superior temporal gyrus, anterior and mid cingulate cortex, as well as right posterior cingulate cortex and superior temporal gyrus. They also observed increased similarity weights in the bilateral lateral occipital cortex and postcentral gyri, left orbitofrontal cortex, and right dorsolateral prefrontal cortex. Thus, frontotemporal alterations emerge consistently across studies, representing a potential biological marker for BD.

Notably, this is the first study to investigate MIND networks in a large, multi-site cohort of controls and subjects with BD. The multi-site nature of our dataset provided a more general representation of the BD population by incorporating the added heterogeneity of retrospective MRI data from various international sites. However, MRI datasets from different sites are not directly comparable, and biological effects of interest could be masked by differences between sites. Multi-site harmonization is the standard approach to address the biases introduced by site differences. Site effects are also known to confound ML and DL models, which tend to capture them at the expense of biological outcomes of interest (*47*). We integrated an innovative multi-site harmonization pipeline into the LOSO-CV framework, effectively harmonizing both the training and independent test sets. In each LOSO-CV iteration, the holdout test set was harmonized a posteriori using the reference-based approach introduced in M-ComBat (*48*). This enabled us to preserve the independence of the test set and prevent data leakage. We evaluated the effectiveness of this harmonization using qualitative assessments of the data UMAP projections before and after harmonization, which demonstrated successful harmonization for all CV iterations (Fig. S3-S8). Importantly, M-ComBat has previously been validated for harmonizing independent sites in multi-site MRI studies (*49, 50*).

Regarding the LOSO-CV results, only one out of four evaluated models demonstrated a generalizability capacity. To disentangle the contributions of graph and model configuration choices from those of harmonization, we evaluated the models under the LOSO-CV without harmonizing the independent test set (Fig. S9). The analysis showed that harmonization had little impact on all models’ performance, except for site 5, where a slight performance degradation was observed for the best model. The latter suggests that graph and model properties partly captured site-related differences and can potentially help mitigate these effects. Notably, site 5 emerged as the most difficult site for the models to generalize, showing that harmonization is essential when an independent site differs greatly from the rest of the data. These results underscore the importance of evaluating DL models in multi-site settings, which is crucial if research-based diagnostic models are to be translated into clinical practice. This innovative harmonization framework, which works at the post-deployment testing stage of models, is a step further in this direction.

### Limitations and future perspectives

Our study is not without limitations. First, the StratiBip dataset was derived from cross-sectional retrospective data collected from various international sites, which affected the final dataset balancing in terms of numerosity and site-specific information. To balance the entire dataset for age, we downsampled our data, stratifying by site. This led to the exclusion of approximately 200 subjects. An alternative strategy would have been to use upsampling and avoid losing data samples. However, this would have required carefully inspecting the added observations to ensure that no spurious or noisy samples were introduced in the dataset, which was difficult to verify.

In this study, we tested the potential of the implemented DL framework by evaluating only one clinical outcome: diagnosis. However, other clinical variables should be considered that might increase the diagnostic accuracy or influence the model’s explainability, such as information about medication, symptom severity, age of onset, duration of illness, and type of BD, which are known to impact several brain regions. For instance, the basal ganglia are greatly impacted by cumulative use of antipsychotics, and findings associated with differences in these regions might have reflected this factor and not directly to BD pathophysiology. Additionally, a relationship between age and duration of illness could be hypothesized, which we have not disentangled in this study. Regarding the GNN models used, we only tested three standard categories of architectures: GCN, GAT, and GIN. It is possible that more advanced and optimized architectures could have further improved our results. Future work may focus on state-of-the-art DL models, including other GNNs and transformer-based architectures.

We performed a set of sensitivity analyses and validated the resulting models using a LOSO-CV framework. It is worth noting that the sensitivity analyses aimed to reduce the search space of hyperparameters, in terms of both graph design and GNN architectures, rather than selecting the final configurations absolutely. However, we used the entire dataset for this analysis, splitting and stratifying by site. This introduced a certain level of site-based data leakage into the LOSO-CV.

Graph creation was a crucial step that required a sparsification process. We employed a simple threshold-based approach in which non-informative or irrelevant edges were removed from the graph. More advanced sparsification approaches could have been implemented, such as edge attention mechanisms in the GNN. However, this would have substantially increased the computational resources needed for the GNN analysis.

Finally, in terms of model explainability, we merely aggregated the results of all LOSO-CV iterations. Future works should evaluate the expressiveness of this aggregated explainability at the level of the individual models or the consistency of the learned decision functions of the different models in the LOSO-CV.

## Conclusion

In this study, we proposed a robust multi-site graph DL framework based on brain morphometric similarity networks to advance the diagnosis of BD, a disabling disorder that still lacks robust brain markers. For the first time, we evaluated the potential of MIND networks for discriminating BD from controls using GNN analysis, surpassing the diagnostic accuracy of the state-of-the-art literature in a LOSO-CV framework. Using a comprehensive explainability approach, we identified the most important nodes and node features for the model’s decision-making process. Top features mainly involved age, GMV, and MIND similarity profiles in the basal ganglia, frontal lobe, and temporal lobe. Across the graph nodes, we found an interaction among age, regional GMV, and MIND similarity that improved diagnostic accuracy even in age-balanced BD and HC samples. This supports evidence that BD is characterized by altered age effects on brain networks.

Our study contributes to the broader effort of characterizing BD in terms of macroscopic brain structural alterations by unveiling new predictive network patterns that support a neurodevelopmental or brain-aging hypothesis of BD.

## MATERIALS AND METHODS

### Study design

This was a retrospective cross-sectional analysis of 3T T1-weighted sMRI data from participants with BD and HC provided by the StratiBip network(*14*). The study aimed to test the effectiveness of a new GNN framework based on GMV similarity profiles in classifying BD, and to identify candidate brain network markers of disease. The study protocol received ethical approval by the research ethics committees of each site. All participants provided their written informed consent prior to study inclusion.

### Dataset Description

The subset of data used was collected across seven sites from 1106 subjects aged between 16 and 84 years (Table 2). The cohort was composed of 605 HC (median age: 31, 25th–95th percentile: 25–66, range: 18–84 years, 337 females, 268 males) and 501 subjects with BD (median age: 41, 25th–95th percentile: 31–63, range: 16-76 years, 290 females, 211 males). All subjects were recruited and assessed at each site after signing a written and informed consent approved by the competent Ethical Committees, in accordance with the principles in the Declaration of Helsinki. sMRI data acquisition and preprocessing details can be found in Table S8.

**Table 2.** Dataset Demographics.

| Site ID | Institution | Subjects Numerosity | Age (mean ±std, years) |
| --- | --- | --- | --- |
| 1 | University of Verona, Italy | 113 (93 HC, 20 BD) | 33.85±11.72 |
| 2 | Fondazione IRCCS Santa Lucia, Rome, Italy | 450 (250 HC, 200 BD) | 45.04 ± 14.58 |
| 3 | Jena University Hospital, Jena, Germany | 134 (111 HC, 23 BD) | 30.33 ± 9.10 |
| 4 | Fondazione IRCCS Ca' Granda Ospedale Maggiore Policlinico, Milan, Italy | 38 (26 HC, 12 BD) | 30.55 ± 9.07 |
| 5 | IRCCS Ospedale San Raffaele, Milan, Italy | 200 (67 HC, 133 BD) | 41.87 ± 12.79 |
| 6 | University of Pittsburgh School of Medicine, Pittsburgh, PA, USA | 86 (28 HC, 58 BD) | 34.01 ± 7.80 |
| 7 | University of British Columbia, Vancouver, Canada | 85 (30 HC, 55 BD) | 22.72 ± 4.21 |

### Morphometric Similarity Networks

#### Morphometric Inverse Divergency (MIND)

After MRI preprocessing, voxel-based estimates of brain tissue volumes are generated, and each ROI is associated with a distribution of voxel intensity values. To estimate the pairwise morphometric similarity between ROIs, we used the MIND algorithm (*19*), which differs from distance metrics such as absolute difference or correlation-based distance (*1, 51*). MIND uses the Kullback–Leibler (KL) divergence, estimated with a non-parametric nearest-neighbour approach (*52*), to compute a similarity score between two regions based on the distribution of sMRI features in the relative vertices or voxels (for surface-based and volume-based approaches, respectively). In this study, MIND was computed in a univariate fashion, using the voxel-based GMV maps for a given brain parcellation, resulting in pairwise MIND scores between ROIs. For each pair of regions, 𝒂 (with 𝒏 voxels) and 𝒃 (with 𝒎 voxels), the algorithm computes: i) the Euclidean distance 𝒓_𝒊_from each sample in 𝒂 to its second-nearest neighbour within 𝒂 (to avoid self-match), and ii) the Euclidean distance 𝒔_𝒊_ to the nearest neighbour of that sample in 𝒃. The estimation of the KL divergence is then computed as:

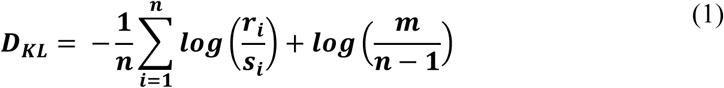

To obtain a symmetric similarity measure, the divergence is estimated as the sum of the divergence of 𝒂 with respect to 𝒃 and the divergence of 𝒃 with respect to 𝒂:

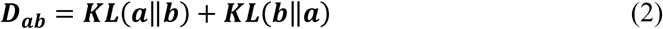

Finally, the MIND score is calculated as:

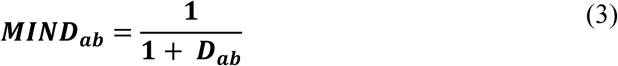

Where 𝑴𝑰𝑵𝑫_𝒂𝒃_is bounded between 0 and 1, the latter indicating maximal similarity between two regions, for consistency with standard correlation-based connectivity metrics. The calculation of the MIND score is repeated for all possible combinations of 𝑵 ROIs, resulting in 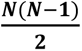 pairwise similarity values, which fill the upper and lower triangle of a symmetric 𝑵 x 𝑵 morphometric similarity matrix used to represent the MIND networks, also denominated the graph adjacency matrix, 𝑨.

We used the voxel-based GMV maps and the neuromorphometrics brain atlas parcellation (*20*) to extract GMV distributions for each ROI and in turn the ROI-to-ROI MIND scores. This resulted in an undirected, weighted, fully-connected graph adjacency matrix of morphometric similarity of dimension 122×122, per subject.

#### Graph Construction

We construct the final brain graphs, 𝑮, from the graph adjacency matrix, 𝑨 ∈ ℝ^𝑵×^ ^𝑵^ . Each subject graph 𝑮_𝒋_will be represented by a set of nodes 𝑽, and edges, which vary for each subject, 𝑬_𝒋_, 𝑮_𝒋_(𝑽, 𝑬_𝒋_). Each node 𝒏_𝒊_ has associated node features 𝑿, represented by a matrix 𝒙_𝒊_ ∈ ℝ^𝑵×𝒅^, where 𝒅 denotes the dimension of the feature set, and 𝑵 the total number of nodes. On the other hand, the edges can be binary or have associated edge features, defined as a matrix 𝜺_𝒔_ ∈ ℝ^𝑴𝒋×𝒛^, where 𝑴_𝒋_ is the number of edges for subject 𝒋 and 𝒛 the dimension of the feature set, with 𝒛 = 𝟏 when edges are weighted. In this study, we focus on a graph classification task, where each graph 𝑮_𝒔_ is associated with a label 𝒚_𝒋_. The final graphs are composed of the 122 nodes, corresponding to the 𝑵 ROIs in the neuromorphometrics atlas, and an edge is defined between two nodes if a pairwise connection exists. In principle, MIND networks are fully connected, and a sparsification approach is applied to reduce them to sparse, interpretable graphs. The standard methodology is to threshold the adjacency matrix 𝑨, keeping only the top 𝒌 percentage of connections (*53*), as low magnitude connections are considered to be spurious. The initial graph density threshold is set to top-𝟑𝟎%, which is deemed as a sufficient sparsity level (*54, 55*), and the edges are defined as binary, i.e., without edge features. The node features were initially set to the MIND values, where 𝒙_𝒊_ represents the node 𝒏_𝒊_ similarity profile and 𝒅 = 𝟏𝟐𝟐. The graph density and node feature combinations were tuned in the sensitivity analysis.

### Graph Convolutional Neural Networks

GNN have emerged as suitable DL models to analyse non-Euclidean data, such as graphs. GNNs main operating principle is the message-passing mechanism. Input graphs are processed by considering each node’s neighbours. As the depth of the model increases (L), the receptive field expands accordingly, covering a k-hop distance from the target node. The nodes transport node feature information which are transformed by the model into updated embeddings, which are then aggregated at graph-level to produce output predictions for a given task. More details on GNNs can be found in Supplementary Materials.

In this study, three GNN architectures were compared: GCNs, GATs, GINs. GCN architecture was introduced by Kipf and Welling in 2017 (*56*). In the convolutional layer, the message transformation is linear, given by the message computation 𝒎_𝒖→𝒗_ = 𝑾𝒙^𝑳^, where 𝑾 is the matrix of learnable weights, and 𝒖 is a neighbouring node of 𝒗. The message is then normalized by the degree of the nodes involved and aggregated across all k-hop neighbouring nodes. Thus, in a GCN, the neighbour and message importance are determined by the graph topology as the message is weighted by the node’s degree. Finally, a non-linear activation is applied to the neighbours’ aggregated messages. The model architecture consists of *L* convolutional layers, each with *d* hidden features, which determines the new node embedding dimension, followed by a fully connected layer (or a global pooling layer) to map the final embeddings to the specific output dimensions required for the classification task. For details on GIN and GATs, see Supplementary Materials.

### Models Training

All GNN models were trained using cross entropy loss and the Adam optimiser with a learning rate of 0.001, evaluated over a maximum of 100 epochs. An early stopping strategy was implemented, interrupting the training process after 20 consecutive epochs with no improvement in the validation loss. The experimental framework was implemented in Python using the PyTorch and PyTorch Geometric libraries, with computation performed on NVIDIA GPUs provided via the Google Colab environment. For the ML models, standard architectures from sklearn library were used.

### Model Validation Framework: LOSO-CV

In the LOSO-CV, we evaluated the model’s generalizability across different independent sites, as shown in Fig. 5. The internal 5-fold CV was used to select the best model configuration among different hyperparameter options. Age balancing was applied within each fold to the training set as well as the multi-site data harmonization, which was applied both at the 5 internal folds of the CV using ComBat (*21, 22*), as well as in the outer fold of the LOSO-CV using M-ComBat (*48, 49*), to control for site effects in training, validation, and external test sets. Details about harmonization can be found in Supplementary Materials.

**Fig. 5.**
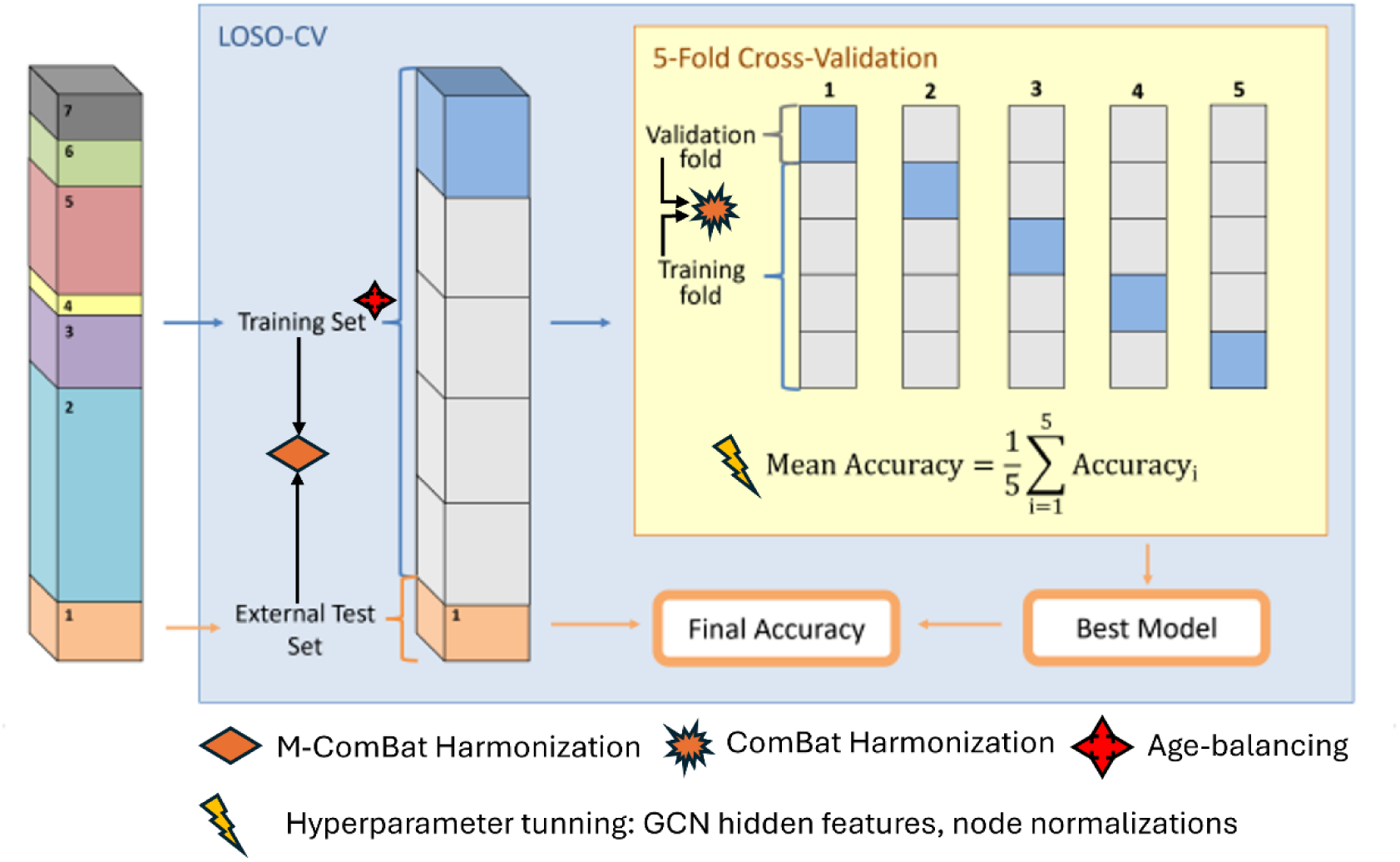
**LOSO-CV pipeline scheme.**

### GNN Explainability

As DL models become increasingly popular in neuroscience, the need for interpretability has become central to ensuring their scientific validity and clinical reliability. Several methods have been developed that extend standard xAI to the GNN field. In this study, we applied two different methods: CAM (*57*) and GNNExplainer (*58*). Both were used to explain the best model from the LOSO-CV framework, the model validation stage. The xAI results were extracted for all folds and then averaged to retrieve global node and feature importance scores.

#### Class Activation Mapping

CAM is a technique originally developed for convolutional neural networks in image classification tasks and later adapted to GNNs (*59*) to reflect the graph nodes’ contributions to a classification task. CAM takes as input the final node embedding, 𝑒_𝑛_, before the global average pooling (GAP) operation, and the row 𝑤^𝑐^ of the classification weight matrix corresponding to a given predicted class 𝑐. The node 𝑛 contribution to the output score 𝑦^𝑐^ for the predicted class is computed as:

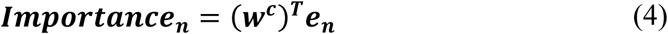

The process was repeated for each graph in the external test batch, and the resulting node-level importance scores were aggregated across samples. The main drawback of CAM is the inability to disentangle node importance from node feature importance, limiting its granularity.

#### GNNExplainer

GNNExplainer is a widely used method for GNN explainability, introduced to identify which parts of a graph are most influential for a given prediction. GNNExplainer works by learning a mask over the node input features, represented by a subgraph of 𝐺, denoted as 𝐺_𝑠_, and a subset of node features 𝑋, denoted as 𝑋_𝑠_, that maximally preserves the model’s prediction 𝑌, identifying the most reliable subset of information. Thus, the GNNExplainer optimization framework is formulated as the maximization of the mutual information (𝑀𝐼) between predicted label 𝑌 and explanation (𝐺_𝑠_, 𝑋_𝑠_):

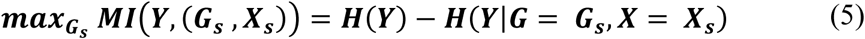

Where 𝑀𝐼 quantifies the change in probability of a prediction when the model sees a restricted computation graph, i.e., limited to 𝐺_𝑠_and 𝑋_𝑠_. 𝐻(𝑌) is a fixed-term entropy given by the original GNN trained on 𝐺, and the term 𝐻(𝑌|𝐺 = 𝐺_𝑠_, 𝑋 = 𝑋_𝑠_) represents the conditional entropy for a prediction of 𝑌 based on 𝐺_𝑠_and 𝑋_𝑠_. The difference between these terms quantifies how much 𝐺_𝑠_ and 𝑋_𝑠_would change prediction 𝑌, thus the GNNExplainer objective entails the minimization of the term 𝐻(𝑌|𝐺 = 𝐺_𝑠_, 𝑋 = 𝑋_𝑠_). For more details, see the original article.

## Data Availability

Data associated with this study is not publicly available.

## List of Supplementary Materials

Results

Fig. S1 to Fig S10

Table S1 to Tables S7

Materials and Methods

Fig. S11

Table S8 References (*1–16*)

## Acknowledgments

EM was supported by the European Union – NextGeneration EU (PRIN 2022 PNRR, grant n. P20229MFRC).

PB was partially supported by the Italian Ministry of Education and Research – MUR (‘Dipartimenti di Eccellenza’ Programme 2023-27 - Dept. of Pathophysiology and Transplantation, Universita degli Studi di Milano), the Italian Ministry of Health (Hub Life Science - Diagnostica Avanzata, HLS-DA, PNC-E3-2022-23683266 - CUP: C43C22001630001 / MI-0117; RC 2026), and by the ERANET Neuron JTC 2023 (ERP-2023-23684211 - ResilNet) and Eranet Neuron JTC 2024 (ER-2024-23684536 - BRAWO Project).

## Author contributions

Conceptualization: EM, PB

Methodology: GP, IWS, EM

Formal analysis: GP

Visualization: IWS

Funding acquisition: PB, MB, FB, IN, MLP, FP, JCS, YL

Project administration: PB, JCS

Supervision: EM, PB

Writing – original draft: IWS, AP, EM

Writing – review & editing: EM, AP, GP, AMB, YT, PB, JCS

## Data and materials availability

All code is publicly available in GitHub https://github.com/GiuseppePoli/MBNGNN.git. Data associated with this study is not publicly available. A request to access the data should be send to PB or JCS.

