## Supplementary Materials for "Advancing diagnosis of bipolar disorder using brain morphometric similarity networks in a graph AI framework"

#### RESULTS

##### Sensitivity Analysis

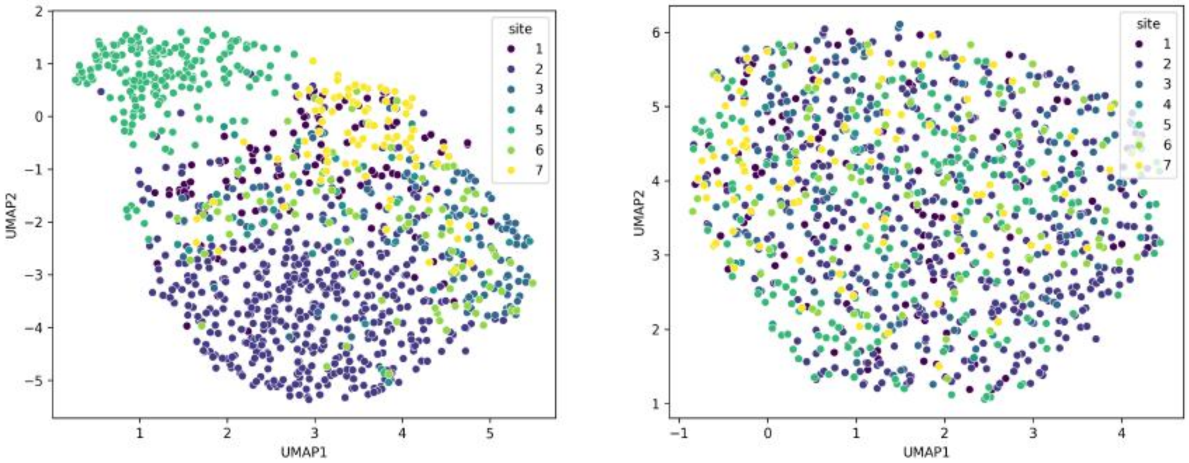

**Fig. S1.** Multi-site harmonization: qualitative evaluation of ComBat harmonization results. The UMAP dataset projection results are presented before (left) and after (right) harmonization. Before harmonization, data clusters-by-site are formed, indicating the presence of site effects in the dataset. After harmonization, the clusters-by-site disappear, suggesting an efficient homogenization of the multi-site data.

**GNN Model Benchmark.** Three GNN architectures were evaluated: i) graph attention networks (GAT)*(1)*, ii) graph isomorphism networks (GIN) *(2)*, and iii) graph convolutional networks (GCN) *(3)*, and compared against two baseline machine learning (ML) models, a support vector machine (SVM) and extreme gradient boosting (XGBoost). Different parameters were compared in the validation set: specifically, for the GNN architecture, the number of layers (2, 3, 4) and hidden dimensions (32, 64, 128); at the graph-level, two different combinations of node features were compared: only MIND values and MIND values plus age, sex and ROI GMV (MIND + all). Here, the MIND networks' graph topology was kept fixed with the top-30% of connections and with binary edges, i.e., no edge features. The model results on the test set are shown in Table S1. When using MIND values alone as node features, the GCN achieved the highest accuracy (63.25%), outperforming both GNNs, GIN at 62.05% and GAT at 58.43%, and ML models. This pattern was still observed when sex, age, and ROI GMV were integrated into the node attributes (MIND + all), with GCN reaching a peak accuracy of 66.27%, compared with GIN at 63.86% and GAT at 62.65%. Notably, performance gains achieved by adding sex, age, and ROI GMVs as features were consistent across architectures, with XGBoost improving the most. The GCN model stood out as the best model due to its superior performance, outperforming all models by 4% accuracy points when only MIND was used. The superior performance of GCN

highlights GNN's ability to exploit graph topology, identifying discriminative information that traditional methods may overlook.
The GCN model was therefore selected for all subsequent sensitivity analyses, considering different numbers of layers (2, 3, 4) and hidden dimensions (32, 64, 128).

**Table S1.** Model test set performances for BD classification. The best performance is highlighted in bold and the second in underlying.

| Models | MIND | MIND+all |
|---|---|---|
| GAT | 58.43 | 62.65 |
| **GCN** | **63.25** | **66.27** |
| GIN | 62.05 | 63.86 |
| SVM | 59.64 | 62.65 |
| XGboost | 59.04 | 64.46 |

**Features Selection**. In the GCN analysis, different combinations of node attributes were evaluated, with a primary focus on integrating demographic and anatomical information with MIND connectivity values. Single and multiple types of features were considered. Single-feature configurations consisted of (i) node-specific MIND similarity profile (MIND values between the ROI and all the others), (ii) node-specific ROI-GMV, and subject-specific (iii) sex and (iv) age, which were repeated across all network nodes. The ROI-GMVs were previously corrected for total intracranial volume (TIV) by normalization. Here, graph density was kept fixed at top-30%, edges were binary, and a GCN model was used in all comparisons (while varying numbers of layers and hidden dimensions, as described in the previous section). Table S2 shows the performances on the test set for single node features and varying combinations. Among the single-feature models, only the one using MIND values provided acceptable results, while the configurations using ROI-GMV, sex, and age resulted in accuracies at the chance level, respectively 54.82% and 56.63%. These results suggest that, although the graph topology is implicitly encoded in the graph through the subject-level edge structure, the single features age, sex, and ROI volumes, in combination with the graph skeleton, did not have sufficient predictive power on their own. Instead, the MIND similarity as a node feature achieved the best performance, even when compared with the standard ROI-GMV. This is further evident when MIND values are combined with other features, reaching the greatest accuracy with age and ROI volumes (68.07%) and a slightly lower one when all node features were combined (66.27%). Interestingly, age information was not predictive on its own, suggesting no differences in graph-by-age relationships between groups. However, in multi-feature configurations, age consistently led to improved performances when combined solely with MIND and with MIND plus sex and/or ROI-GMV. These findings suggest the importance of including node-level morphological similarity data and a relevant integration of it with age, which aids in the distinction between groups. For the subsequent sensitivity analyses, we used the combination of features that yielded the best performance, MIND + age + ROI-GMV, and MIND+all (MIND + age + ROI-GMV + sex) for consistent comparisons between all sensitivity analyses.

**Table S2.** Comparison of node feature combinations for the GCN analysis for discrimination of HCs from the BD group. The best result is highlighted in bold, and the second and third are in underlying.

| Nº of node feature-types | Combination set | Test set accuracy (%) |
|---|---|---|
| 1 | **MIND** | **63.25** |
| | Sex | 54.82 |
| | Age | 54.82 |
| | ROI-GMV | 56.63 |
| 2 | MIND + sex | 60.24 |
| | **MIND + age** | **66.27** |
| | MIND + ROI Volumes | 61.45 |
| | Age + ROI-GMV | 56.02 |
| 3 | <u>MIND + Sex + Age</u> | <u>66.87</u> |
| | MIND + Sex + ROI Volumes | 64.46 |
| | <u>**MIND + Age + ROI Volumes**</u> | <u>**68.07**</u> |
| 4 | **MIND + sex + age + ROI Volumes** | **66.27** |

**Graph density.** The impact of graph density on the GCN model performance was evaluated. We tested top-K thresholds for 10%, 30%, 70% and 90% of edges, where the first retains only the strongest 10% connections, yielding the sparsest graph, and the last retains the strongest 90% connection, resulting in the densest graph. The GCN model was tested with varying layers and hidden dimensions. Two node feature combinations were used: the best performing combination, MIND + age + ROI-GMV, and MIND+all. Table S3 reports the test set performances on the best GCN configurations. For the MIND + age + ROI-GMV combination, the model performance was not affected by the graph density, whereas it varied when sex was added for the MIND+all experiment. In the latter case, the best performance was obtained with a density of 10%, for an accuracy of 68.27%.

**Table S3.** GCN model performance on the test set for varying graph sparsification density levels.

| Density Level | MIND+age+ROI-GMV+sex | MIND+age+ROI-GMV |
|---|---|---|
| 90% | 66.87% | 68.07% |
| 70% | 66.27% | 68.07% |
| 30% | 66.27% | 68.07% |
| **10%** | **68.67%** | 68.07% |

**GCN Hyperparameter Tuning.** The GCN architecture was evaluated through the comparison of performances with varying numbers of layers and hidden features, using MIND+all as a configuration of node features. For each model architecture configuration, different node features normalization techniques were tested: i) dividing by standard deviation (norm_sd), removing the mean (norm_mean), standardizing (norm_std), dividing by the square root of standard deviation (norm_rsd), scaling with p-rooth of the variance (norm_pr_v), and through layer normalization (layernorm). Additionally, the previous four graph densities were tested, resulting in 6x4 trials per GCN configuration. Fig. S2 shows the results (mean and standard deviation across all density levels and node normalization methods) for each GCN configuration. Increasing the GCN's depth degraded its performance, a result expected due to the over-smoothing problem of GNNs. The GCN with 2 layers achieved better average performance when compared to using 3 or 4 layers, with the added computational advantage of being lighter to train. For this reason, a GCN with 2 layers was used for the final model validation stage.

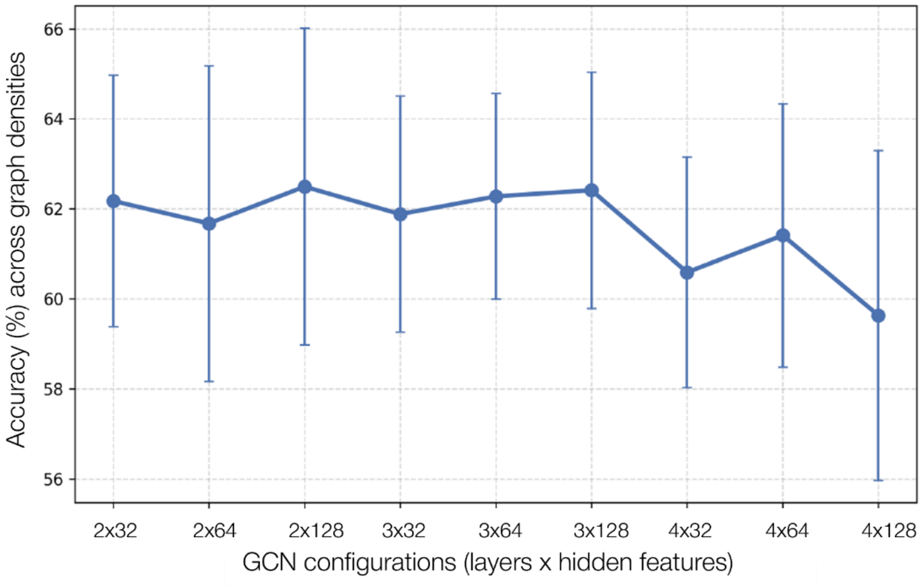

**Fig. S2.** GCN architecture evaluation. For each layer x hidden feature combination, six different node normalization and four graph densities were used, resulting in 24 trials. The mean performance of the GCN model for all trials is shown, with the corresponding standard deviation.

#### Model Validation: Leave-One-Site-Out Cross-Validation

**Multi-site MIND networks Harmonization.** *M*-ComBat *(4)* model was employed to harmonize the test set subjects' MIND networks within the LOSO-CV frameworks, as in *(5)*. Here, the subsets used as training set were harmonized with ComBat, and the external holdout site was harmonized a posteriori, during the model testing phase, with the harmonized training set, employing M-ComBat (detailed pipeline explained in *Multi-site ComBat Harmonisation* in *Methods*). Fig. S3-S8 shows the UMAP data projections for each of the folds of the LOSO-CV, each with site *i* as the holdout test set, for the pre- and post-ComBat harmonization of the training set.

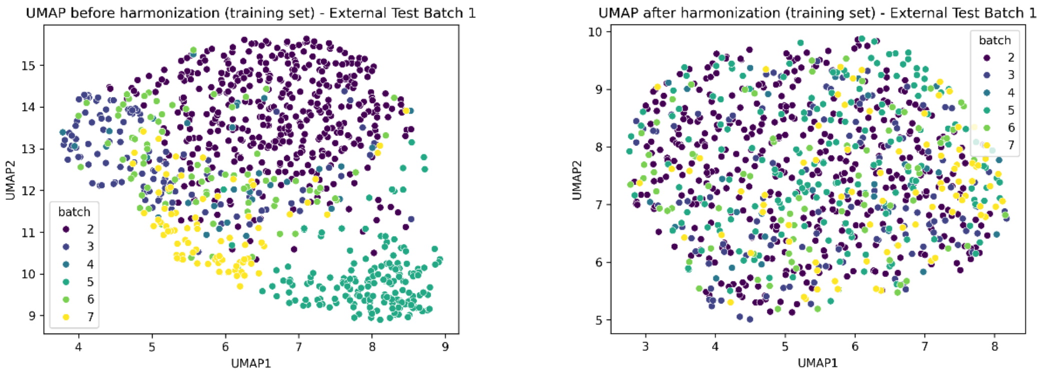

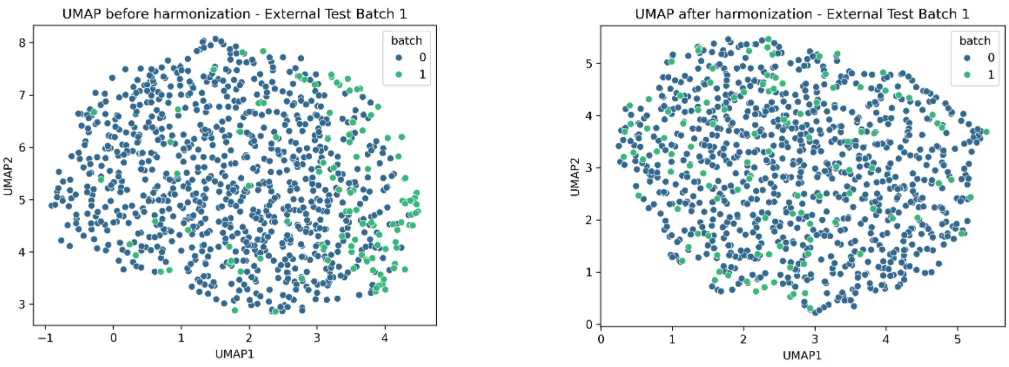

**Figure S3.** Qualitative evaluation of M-ComBat harmonization results for fold n=1. In the top figure, the UMAP projection results for the training set (excluding site 1) ComBat harmonization are presented. In the bottom figure, the UMAP projection results are shown for the M-ComBat harmonization of the independent test set from site 1 (batch=1), with the harmonized training set (represented by batch=0), before (left) and after (right) with the harmonized training set.

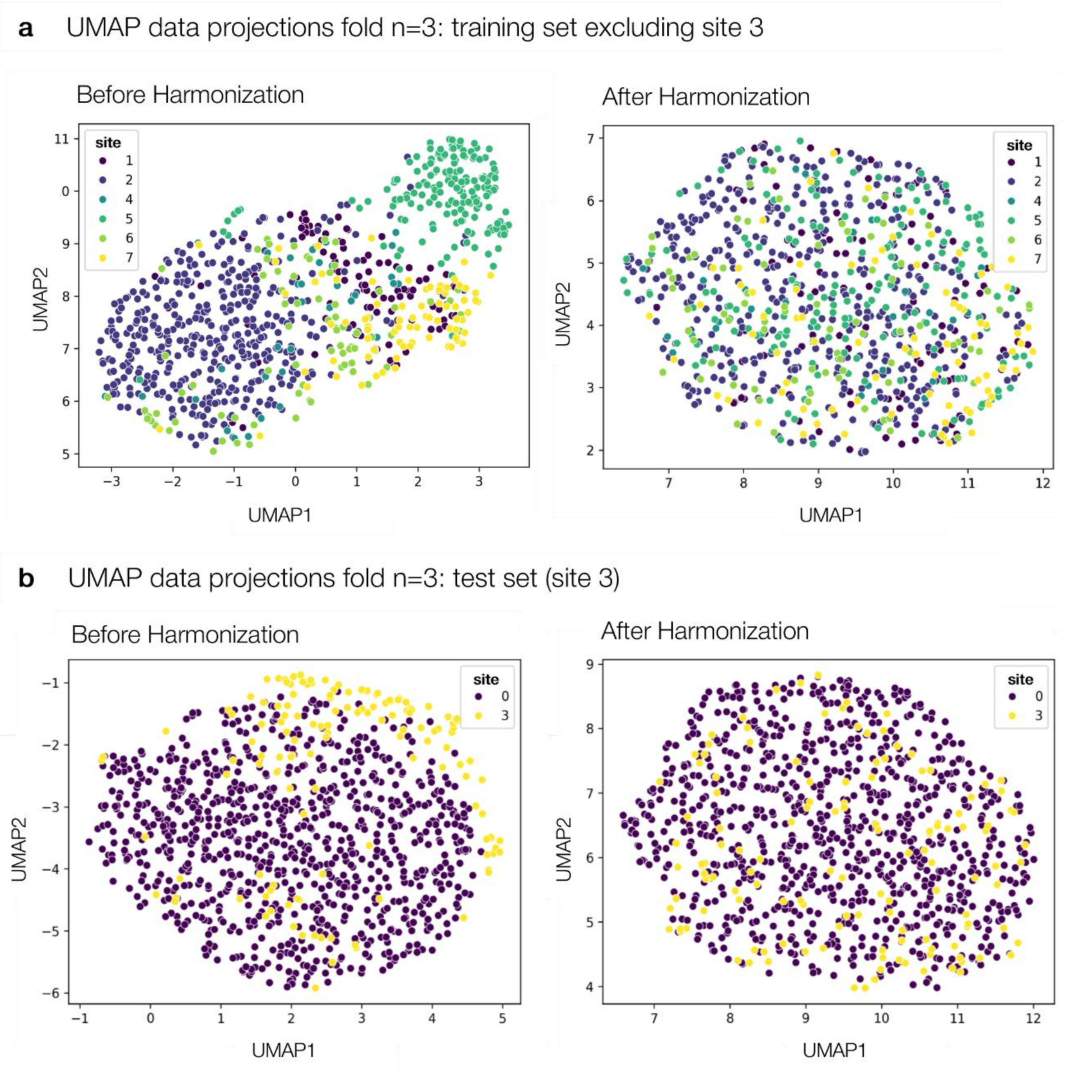

**Fig.S4.** Qualitative evaluation of M-ComBat harmonization results for fold n=2. In the top figure, the UMAP projection results for the training set (excluding site 3) ComBat harmonization are presented. In the bottom figure, the UMAP projection results are shown for the M-ComBat harmonization of the independent test set from site 3 (batch=3), with the harmonized training set (represented by batch=0), before (left) and after (right)  with the harmonized training set.

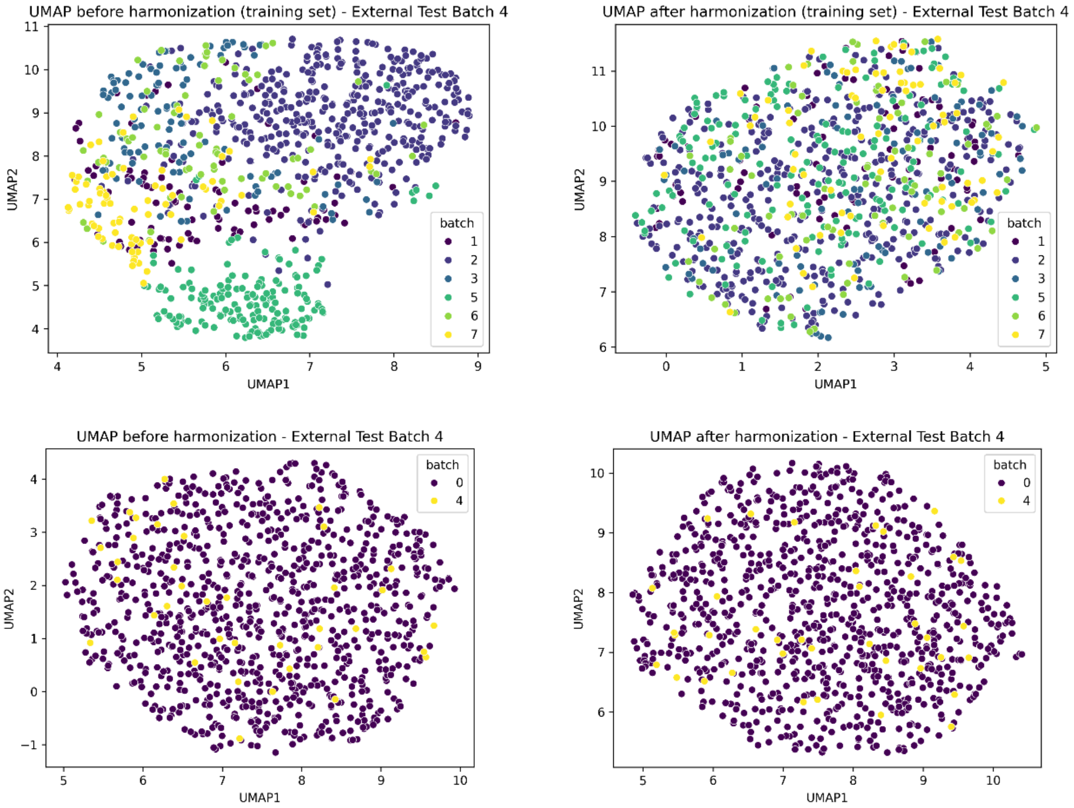

**Fig. S5.** Qualitative evaluation of M-ComBat harmonization results for fold n=3. In the top figure, the UMAP projection results for the training set (excluding site 4) ComBat harmonization are presented. In the bottom figure, the UMAP projection results are shown for the M-ComBat harmonization of the independent test set from site 4 (batch=4), with the harmonized training set (represented by batch=0), before (left) and after (right)  with the harmonized training set.

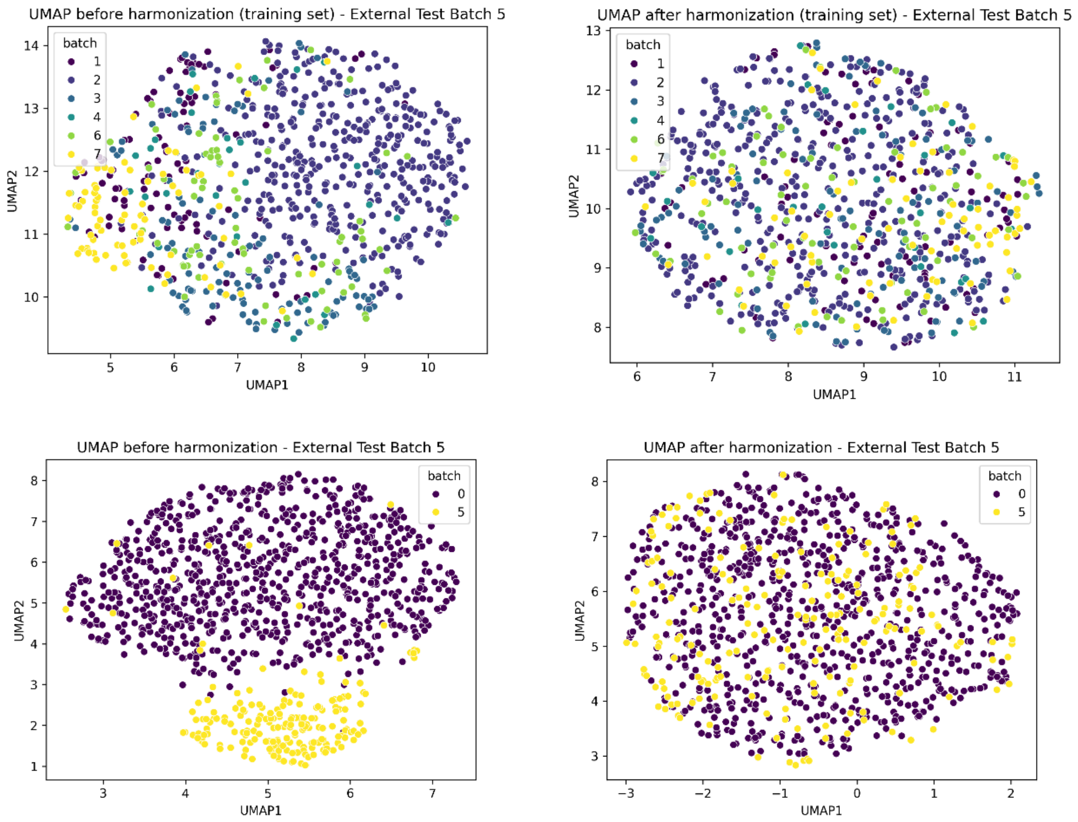

**Fig. S6.** Qualitative evaluation of M-ComBat harmonization results for fold n=4. In the top figure, the UMAP projection results for the training set (excluding site 5) ComBat harmonization are presented. In the bottom figure, the UMAP projection results are shown for the M-ComBat harmonization of the independent test set from site 5 (batch=5), with the harmonized training set (represented by batch=0), before (left) and after (right) with the harmonized training set.

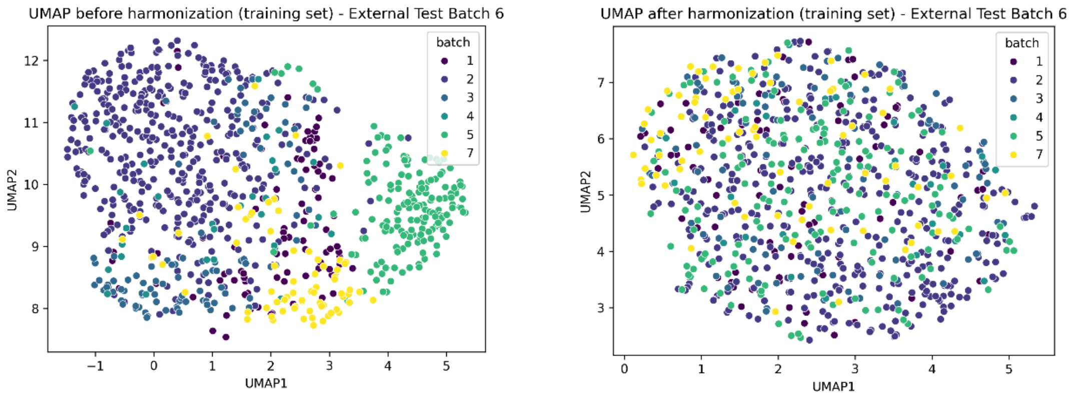

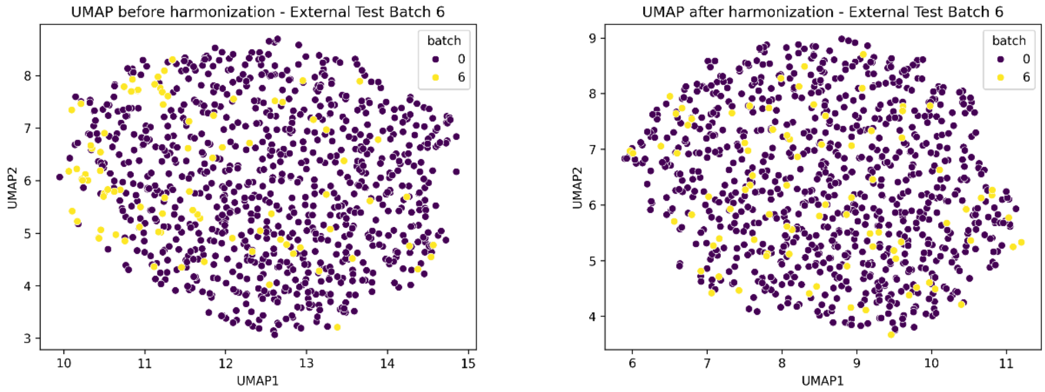

**Fig. S7.** Qualitative evaluation of M-ComBat harmonization results for fold n=5. In the top figure, the UMAP projection results for the training set (excluding site 6) ComBat harmonization are presented. In the bottom figure, the UMAP projection results are shown for the M-ComBat harmonization of the independent test set from site 6 (batch=6), with the harmonized training set (represented by batch=0), before (left) and after (right) with the harmonized training set.

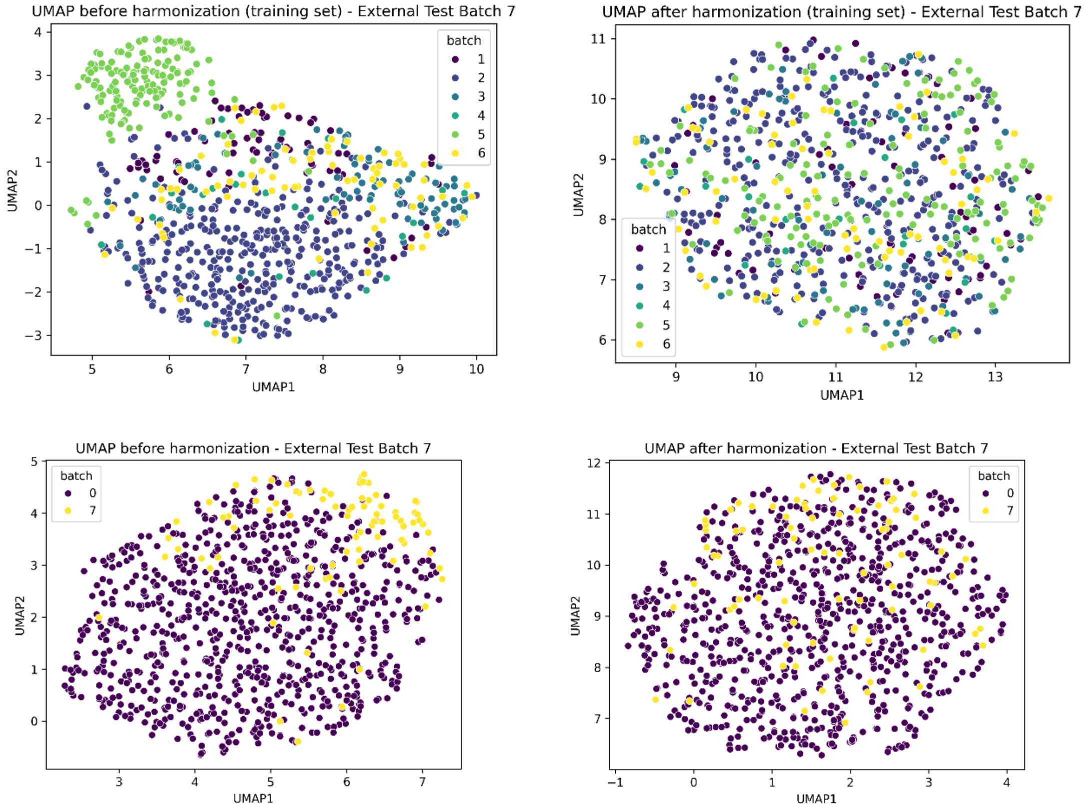

**Fig. S8.** Qualitative evaluation of M-ComBat harmonization results for fold n=6. In the top figure, the UMAP projection results for the training set (excluding site 7) ComBat harmonization are presented. In the bottom figure, the UMAP projection results are shown for the M-ComBat harmonization of the independent test set from site 7 (batch=7), with the harmonized training set (represented by batch=0), before (left) and after (right) with the harmonized training set.

##### *LOSO-CV: Impact of Multi-site Harmonization*

Fig. S9 shows the model performances trained with the harmonized training set but without harmonizing the external test data. The harmonization seems to contribute to the overall model generalizability, with 8 out of 14 trials decreasing their performance without harmonization, although this contribution is almost negligible for all models. Noticeably, OSR ( site 5) seems to benefit from the harmonisation the most, with the highest model performance degradation when harmonization is not performed for the best model. The model difficulty of generalizing to site 5 aligns with findings from the UMAP projections analyses on the raw data, which showed a distinct cluster for this site in the projected feature space, indicating a higher heterogeneity level when compared to the other sites (see Fig. S3-S8). Importantly, the harmonization of the test set was performed fully respecting its independence, where data is harmonized a posteriori in the test phase; therefore, harmonization did not increase the risk for data leakage of any type in our pipeline.

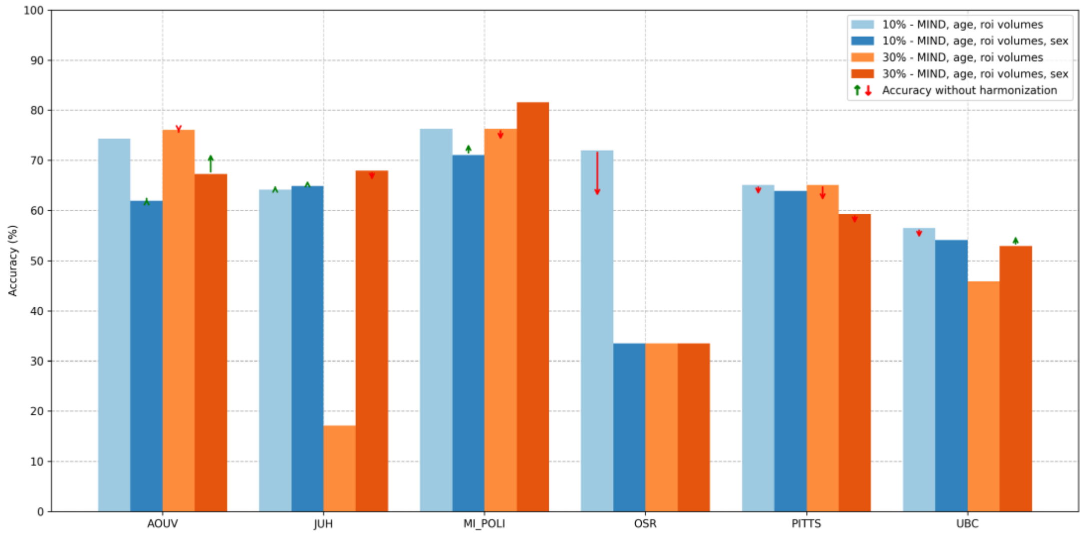

**Fig. S9.** Impact of not harmonizing the independent test sets on model performances. In each fold of the LOSO-CV, the models were trained with the harmonized training set, and applied to the non-harmonized (raw) test sets.

**Comparison with baseline ML.** The performances of the two baseline ML models are shown in Table S4. The models used the MIND similarity values, age, and GMV-ROI as features. These results confirm that the GCN model achieves superior performance on average; however, the Friedman test did not reveal statistically significant differences. It should be noted that the low sample size and high variance across sites limit the power of the test, reducing its reliability. Although the SVM outperformed the GCN for three sites, 1, 3, and 4, it underperformed for 5, 6, and 7, similarly to the XGBoost. The latter indicates that the basic ML models fail to generalize in more difficult out-of-distribution scenarios.

**Table S4.** LOSO-CV accuracy (%) performances and comparison with baseline ML models.

| Model | Site 1 | Site 3 | Site 4 | Site 5 | Site 6 | Site 7 | Average |
|---|---|---|---|---|---|---|---|
| XGBoost | 71.68 | 56.72 | 81.58 | 53.50 | 46.51 | 40.00 | 56.25 |
| SVM | **77.88** | **74.63** | **84.21** | 33.50 | 47.67 | 38.82 | 55.03 |
| GCN | 74.34 | 64.18 | 76.32 | **72.00** | **65.12** | **56.57** | **68.07** |

**Table S5. Neuromorphometric Atlas Labels**

| ID | FeatureName |
|---|---|
| 0 | Right Accumbens Area |
| 1 | Left Accumbens Area |
| 2 | Right Amygdala |
| 3 | Left Amygdala |
| 4 | Right Caudate |
| 5 | Left Caudate |
| 6 | Right Cerebellum Exterior |
| 7 | Left Cerebellum Exterior |
| 8 | Right Hippocampus |
| 9 | Left Hippocampus |
| 10 | Right Pallidum |
| 11 | Left Pallidum |
| 12 | Right Putamen |
| 13 | Left Putamen |
| 14 | Right Thalamus Proper |
| 15 | Left Thalamus Proper |
| 16 | Right Ventral DC |
| 17 | Left Ventral DC |
| 18 | Optic Chiasm |
| 19 | Cerebellar Vermal Lobules I-V |
| 20 | Cerebellar Vermal Lobules VI-VII |
| 21 | Cerebellar Vermal Lobules VIII-X |
| 22 | Left Basal Forebrain |
| 23 | Right Basal Forebrain |
| 24 | Right ACgG anterior cingulate gyrus |
| 25 | Left ACgG anterior cingulate gyrus |
| 26 | Right AIns anterior insula |
| 27 | Left AIns anterior insula |
| 28 | Right AOrG anterior orbital gyrus |
| 29 | Left AOrG anterior orbital gyrus |
| 30 | Right AnG angular gyrus |
| 31 | Left AnG angular gyrus |
| 32 | Right Calc calcarine cortex |
| 33 | Left Calc calcarine cortex |

| | |
|---|---|
| 34 | Right CO central operculum |
| 35 | Left CO central operculum |
| 36 | Right Cun cuneus |
| 37 | Left Cun cuneus |
| 38 | Right Ent entorhinal area |
| 39 | Left Ent entorhinal area |
| 40 | Right FO frontal operculum |
| 41 | Left FO frontal operculum |
| 42 | Right FRP frontal pole |
| 43 | Left FRP frontal pole |
| 44 | Right FuG fusiform gyrus |
| 45 | Left FuG fusiform gyrus |
| 46 | Right GRe gyrus rectus |
| 47 | Left GRe gyrus rectus |
| 48 | Right IOG inferior occipital gyrus |
| 49 | Left IOG inferior occipital gyrus |
| 50 | Right ITG inferior temporal gyrus |
| 51 | Left ITG inferior temporal gyrus |
| 52 | Right LiG lingual gyrus |
| 53 | Left LiG lingual gyrus |
| 54 | Right LOrG lateral orbital gyrus |
| 55 | Left LOrG lateral orbital gyrus |
| 56 | Right MCgG middle cingulate gyrus |
| 57 | Left MCgG middle cingulate gyrus |
| 58 | Right MFC medial frontal cortex |
| 59 | Left MFC medial frontal cortex |
| 60 | Right MFG middle frontal gyrus |
| 61 | Left MFG middle frontal gyrus |
| 62 | Right MOG middle occipital gyrus |
| 63 | Left MOG middle occipital gyrus |
| 64 | Right MOrG medial orbital gyrus |
| 65 | Left MOrG medial orbital gyrus |
| 66 | Right MPoG postcentral gyrus medial segment |
| 67 | Left MPoG postcentral gyrus medial segment |
| 68 | Right MPrG precentral gyrus medial segment |
| 69 | Left MPrG precentral gyrus medial segment |
| 70 | Right MSFG superior frontal gyrus medial segment |
| 71 | Left MSFG superior frontal gyrus medial segment |
| 72 | Right MTG middle temporal gyrus |
| 73 | Left MTG middle temporal gyrus |
| 74 | Right OCP occipital pole |
| 75 | Left OCP occipital pole |
| 76 | Right OFuG occipital fusiform gyrus |
| 77 | Left OFuG occipital fusiform gyrus |

| | |
|---|---|
| 78 | Right OpIFG opercular part of the inferior frontal gyrus |
| 79 | Left OpIFG opercular part of the inferior frontal gyrus |
| 80 | Right OrIFG orbital part of the inferior frontal gyrus |
| 81 | Left OrIFG orbital part of the inferior frontal gyrus |
| 82 | Right PCgG posterior cingulate gyrus |
| 83 | Left PCgG posterior cingulate gyrus |
| 84 | Right PCu precuneus |
| 85 | Left PCu precuneus |
| 86 | Right PHG parahippocampal gyrus |
| 87 | Left PHG parahippocampal gyrus |
| 88 | Right PIns posterior insula |
| 89 | Left PIns posterior insula |
| 90 | Right PO parietal operculum |
| 91 | Left PO parietal operculum |
| 92 | Right PoG postcentral gyrus |
| 93 | Left PoG postcentral gyrus |
| 94 | Right POrG posterior orbital gyrus |
| 95 | Left POrG posterior orbital gyrus |
| 96 | Right PP planum polare |
| 97 | Left PP planum polare |
| 98 | Right PrG precentral gyrus |
| 99 | Left PrG precentral gyrus |
| 100 | Right PT planum temporale |
| 101 | Left PT planum temporale |
| 102 | Right SCA subcallosal area |
| 103 | Left SCA subcallosal area |
| 104 | Right SFG superior frontal gyrus |
| 105 | Left SFG superior frontal gyrus |
| 106 | Right SMC supplementary motor cortex |
| 107 | Left SMC supplementary motor cortex |
| 108 | Right SMG supramarginal gyrus |
| 109 | Left SMG supramarginal gyrus |
| 110 | Right SOG superior occipital gyrus |
| 111 | Left SOG superior occipital gyrus |
| 112 | Right SPL superior parietal lobule |
| 113 | Left SPL superior parietal lobule |
| 114 | Right STG superior temporal gyrus |
| 115 | Left STG superior temporal gyrus |
| 116 | Right TMP temporal pole |
| 117 | Left TMP temporal pole |
| 118 | Right TrIFG triangular part of the inferior frontal gyrus |
| 119 | Left TrIFG triangular part of the inferior frontal gyrus |
| 120 | Right TTG transverse temporal gyrus |

121 Left TTG transverse temporal gyrus

**MIND Similarity Profiles: HC vs BD**

**Brain similarity networks.** The individual-level MIND networks resulted in fully connected 122x122 adjacency matrices (see section *Brain Similarity Networks* in *Methods*). Fig. S10a) shows the resulting distribution of the MIND similarity values across the entire StratiBip dataset for both groups. The average adjacency matrices for HC and BD are shown in Fig. S10b), with their mean difference shown in Fig.S10c). We compared the two groups in terms of their average whole-brain MIND values (averaged over all ROI-to-ROI links) with a Mann-Whitney U-test and found a statistically significant difference (p<0.05, uncorrected), Table S6. When comparing the ROI-specific mean MIND similarity profile between the two groups, 17 ROIs were identified to be statistically significantly different between the two groups (p<0.05, FDR corrected), Table S7.

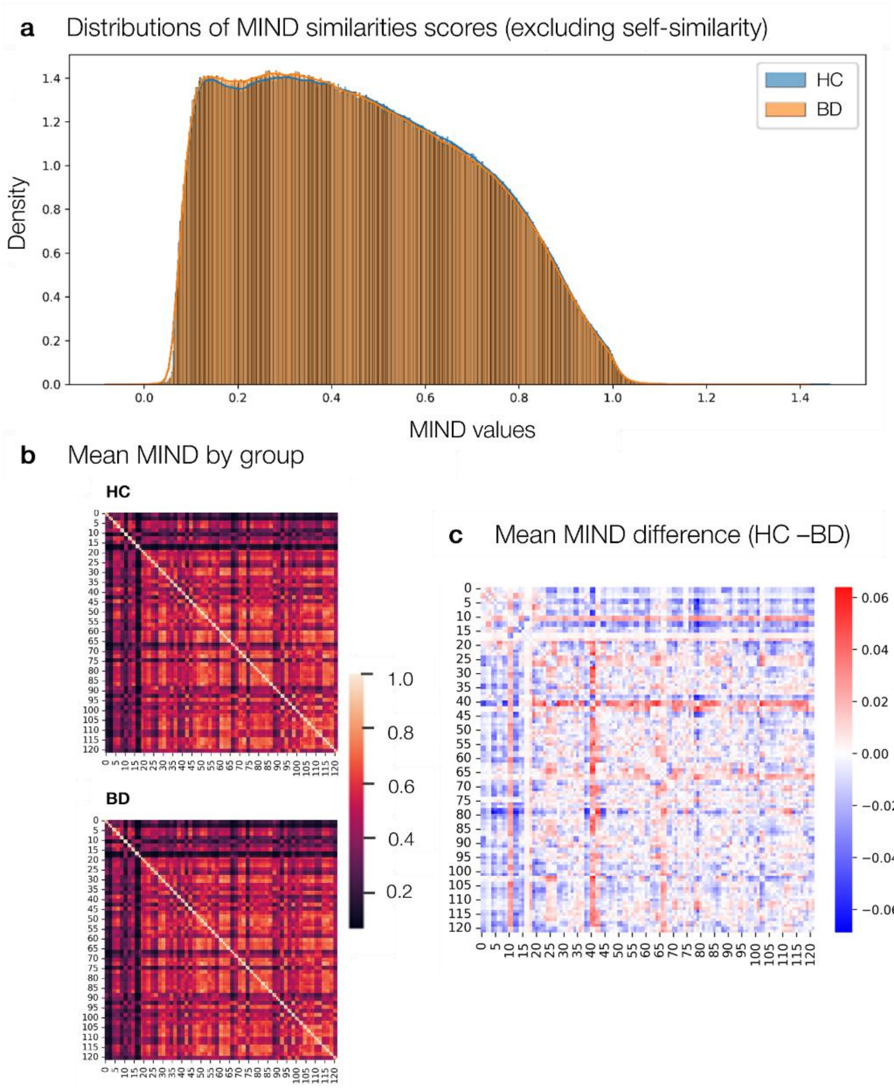

**Fig. S10.** Visual MIND values inspection in the StratiBip sample. a) MIND values distribution for HCs and BD groups. b) The two group averages of the MIND adjacency matrices are presented, on top for the HC group, and on the bottom for the BD group. c) The difference between the average MIND adjacency of the two groups, HC and BD, represented in b). Labels from 0-122 represent the 122 ROIs from the neuromorphometrics brain atlas parcellation, Table S5.

**Table S6.** MIND similarity profiles global comparison. For each subject, the similarity profiles for each brain region-of-interest (ROI), i.e., the similarity of the current ROI with all the others, were averaged to obtain a mean similarity profile. A Mann-Whitney U-Test was used to compare the distributions between the two groups (HC, BD).

| n_HC | n_BD | mean_HC | mean_BD | U | p_uncorrected |
|---|---|---|---|---|---|
| 605 | 501 | 0.46035621 | 0.457670246 | 162710 | 0.034864533 |

**Table S7.** ROI by ROI MIND similarity profiles comparison. For each ROI, the distribution of the similarity profiles for the two groups was compared using a Mann-Whitney U-Test and an FDR correction for multiple comparisons.

| Name | ROI | mean_HC | mean_BD | U | p_uncorrected | p_fdr_bh | significant_fdr | CAM | GNN_ Explainer |
|---|---|---|---|---|---|---|---|---|---|
| Left Putamen | 13 | 0.384424061 | 0.361399607 | 175388 | 6.55969E-06 | 0.000406246 | TRUE | | |
| Right Thalamus Proper | 14 | 0.383508696 | 0.36108337 | 175371 | 6.65978E-06 | 0.000406246 | TRUE | | |
| Right Putamen | 12 | 0.235945408 | 0.254592098 | 128235 | 1.03592E-05 | 0.000421275 | TRUE | | |
| Cerebellar Vermal Lobules I-V | 19 | 0.182165814 | 0.191625872 | 128820 | 1.71644E-05 | 0.000523513 | TRUE | | |
| Left Pallidum | 11 | 0.25043583 | 0.267404806 | 131261 | 0.000124404 | 0.002168177 | TRUE | | |
| Right Amygdala | 2 | 0.249624884 | 0.237365113 | 172012 | 0.000109259 | 0.002168177 | TRUE | | |
| Right frontal pole | 42 | 0.436928374 | 0.454374321 | 131161 | 0.000115166 | 0.002168177 | TRUE | | |
| Left Accumbens Area | 1 | 0.221432235 | 0.209143728 | 171651 | 0.000144239 | 0.002199651 | TRUE | | |
| Left frontal operculum | 41 | 0.447170612 | 0.46372534 | 131901 | 0.000202202 | 0.002740954 | TRUE | | |
| Left Caudate | 5 | 0.43842062 | 0.421090536 | 170296 | 0.000393305 | 0.004798322 | TRUE | | |
| Right orbital part of the inferior frontal gyrus | 80 | 0.545376624 | 0.533016354 | 168018 | 0.001847338 | 0.020488653 | TRUE | | |
| Right Cerebellum Exterior | 6 | 0.449143559 | 0.433184384 | 167866 | 0.002035573 | 0.020694997 | TRUE | | |
| Left entorhinal area | 39 | 0.392966185 | 0.376601708 | 167475 | 0.002603465 | 0.024432516 | TRUE | | |

| | | | | | | | |
|---|---|---|---|---|---|---|---|
| Left Hippocampus | 9 | 0.324011567 | 0.309632576 | 166528 | 0.004626366 | 0.035422353 | TRUE |
| Right precentral gyrus | 98 | 0.41577525 | 0.407189928 | 166553 | 0.004558418 | 0.035422353 | TRUE |
| Right Ventral DC | 16 | 0.425172208 | 0.413914012 | 166521 | 0.004645555 | 0.035422353 | TRUE |
| Left frontal pole | 43 | 0.451698804 | 0.46317937 | 136803 | 0.005283509 | 0.037916946 | TRUE |
| | 121 | 0.39840055 | 0.390164094 | 165378 | 0.008936327 | 0.057635251 | FALSE |
| | 86 | 0.562395197 | 0.556793744 | 165370 | 0.008975982 | 0.057635251 | FALSE |
| | 61 | 0.582720391 | 0.576673711 | 165221 | 0.009743838 | 0.05943741 | FALSE |
| | 62 | 0.588782432 | 0.583204237 | 165096 | 0.010432608 | 0.060608483 | FALSE |
| | 44 | 0.424833323 | 0.435650305 | 138280 | 0.012076492 | 0.06696964 | FALSE |
| | 45 | 0.469736233 | 0.459167259 | 164260 | 0.016258845 | 0.086242567 | FALSE |
| | 17 | 0.146406752 | 0.148059608 | 139489 | 0.022532816 | 0.114541814 | FALSE |
| | 10 | 0.306902169 | 0.297538534 | 163340 | 0.025809302 | 0.125949395 | FALSE |
| | 15 | 0.450878448 | 0.441592446 | 163004 | 0.030347321 | 0.14239897 | FALSE |
| | 109 | 0.544523701 | 0.539274718 | 162393 | 0.040366419 | 0.15886139 | FALSE |
| | 101 | 0.457371034 | 0.451240088 | 162525 | 0.037991866 | 0.15886139 | FALSE |
| | 107 | 0.526385802 | 0.518802529 | 162495 | 0.038520841 | 0.15886139 | FALSE |
| | 67 | 0.283192503 | 0.295332799 | 140489 | 0.036424946 | 0.15886139 | FALSE |
| | 21 | 0.462215809 | 0.452799406 | 162421 | 0.039852472 | 0.15886139 | FALSE |
| | 122 | 0.407827074 | 0.400129614 | 162322 | 0.041694893 | 0.158961779 | FALSE |
| | 100 | 0.472668242 | 0.466044287 | 162009 | 0.048000904 | 0.177457888 | FALSE |
| | 99 | 0.465783997 | 0.458594611 | 161746 | 0.053901932 | 0.190113076 | FALSE |
| | 18 | 0.144936246 | 0.14642611 | 141386 | 0.054540636 | 0.190113076 | FALSE |
| | 30 | 0.534097645 | 0.527859878 | 161607 | 0.05725786 | 0.194040526 | FALSE |
| | 65 | 0.551285666 | 0.556109667 | 141735 | 0.063379827 | 0.203482602 | FALSE |
| | 3 | 0.17041581 | 0.166153884 | 161391 | 0.062816389 | 0.203482602 | FALSE |
| | 31 | 0.570780498 | 0.566677112 | 161025 | 0.073249382 | 0.229139093 | FALSE |
| | 8 | 0.468881412 | 0.461571777 | 160897 | 0.077216956 | 0.235511716 | FALSE |
| | 85 | 0.564586694 | 0.560401824 | 160788 | 0.080732085 | 0.240227181 | FALSE |
| | 40 | 0.398570658 | 0.388912009 | 160364 | 0.095659581 | 0.277868307 | FALSE |
| | 68 | 0.301671318 | 0.312101319 | 142847 | 0.099717564 | 0.282037173 | FALSE |
| | 23 | 0.525779932 | 0.519513551 | 160207 | 0.101718325 | 0.282037173 | FALSE |
| | 108 | 0.521610866 | 0.514069452 | 160118 | 0.105286278 | 0.285442798 | FALSE |
| | 106 | 0.520904041 | 0.515925694 | 159803 | 0.11871777 | 0.314860173 | FALSE |
| | 82 | 0.530689681 | 0.526043392 | 159676 | 0.124498915 | 0.318324846 | FALSE |
| | 72 | 0.544045185 | 0.539860567 | 159660 | 0.125242563 | 0.318324846 | FALSE |
| | 51 | 0.570150762 | 0.564956453 | 159516 | 0.132092094 | 0.328882356 | FALSE |
| | 77 | 0.530618976 | 0.525643903 | 159373 | 0.139177778 | 0.335519354 | FALSE |
| | 120 | 0.556699207 | 0.550467408 | 159193 | 0.148508567 | 0.335519354 | FALSE |
| | 115 | 0.557090269 | 0.554007218 | 159272 | 0.144356189 | 0.335519354 | FALSE |
| | 119 | 0.546163331 | 0.539821158 | 159285 | 0.143681496 | 0.335519354 | FALSE |
| | 20 | 0.507047458 | 0.499003732 | 159208 | 0.147713203 | 0.335519354 | FALSE |

| | | | | | | |
|---|---|---|---|---|---|---|
| 66 | 0.548983778 | 0.552533281 | 144077 | 0.157474673 | 0.343069823 | FALSE |
| 70 | 0.424890855 | 0.41728756 | 159054 | 0.156035225 | 0.343069823 | FALSE |
| 102 | 0.456283126 | 0.450833336 | 158780 | 0.171714302 | 0.367528857 | FALSE |
| 46 | 0.45713074 | 0.450628351 | 158495 | 0.189243927 | 0.398064811 | FALSE |
| 35 | 0.448640693 | 0.445382002 | 158325 | 0.200309626 | 0.414199565 | FALSE |
| 49 | 0.552725066 | 0.549597357 | 158083 | 0.216865973 | 0.429305588 | FALSE |
| 74 | 0.564400921 | 0.562378158 | 158085 | 0.216725225 | 0.429305588 | FALSE |
| 91 | 0.40989908 | 0.406269044 | 158049 | 0.219268755 | 0.429305588 | FALSE |
| 78 | 0.541781603 | 0.538025547 | 158015 | 0.22169059 | 0.429305588 | FALSE |
| 48 | 0.480781006 | 0.476518649 | 157924 | 0.228266691 | 0.43513338 | FALSE |
| 114 | 0.463625974 | 0.467579036 | 145269 | 0.234757021 | 0.440620871 | FALSE |
| 32 | 0.573867251 | 0.571791559 | 157433 | 0.266146075 | 0.491966986 | FALSE |
| 7 | 0.462938496 | 0.458658577 | 157319 | 0.275526968 | 0.495009154 | FALSE |
| 22 | 0.480134426 | 0.475677356 | 157279 | 0.278871349 | 0.495009154 | FALSE |
| 81 | 0.534249003 | 0.52912696 | 157266 | 0.279964193 | 0.495009154 | FALSE |
| 79 | 0.536358519 | 0.533529945 | 156991 | 0.303764605 | 0.51422815 | FALSE |
| 69 | 0.424635075 | 0.420055206 | 157019 | 0.301281582 | 0.51422815 | FALSE |
| 76 | 0.326799677 | 0.334065073 | 146140 | 0.30608241 | 0.51422815 | FALSE |
| 25 | 0.43603052 | 0.439746271 | 146158 | 0.307693893 | 0.51422815 | FALSE |
| 53 | 0.553005656 | 0.549132449 | 156818 | 0.319406926 | 0.526589797 | FALSE |
| 52 | 0.554647856 | 0.55051505 | 156424 | 0.356963221 | 0.580660172 | FALSE |
| 93 | 0.392272233 | 0.388590017 | 156241 | 0.37531613 | 0.602481157 | FALSE |
| 57 | 0.50689135 | 0.503608101 | 155956 | 0.405035161 | 0.641265885 | FALSE |
| 64 | 0.549839983 | 0.547445454 | 155861 | 0.415245942 | 0.641265885 | FALSE |
| 87 | 0.548496086 | 0.545091938 | 155869 | 0.414380266 | 0.641265885 | FALSE |
| 110 | 0.56454946 | 0.561747109 | 155742 | 0.42824855 | 0.653079039 | FALSE |
| 111 | 0.473557215 | 0.469933546 | 155691 | 0.433892838 | 0.653517607 | FALSE |
| 90 | 0.392486312 | 0.39021426 | 155548 | 0.449946342 | 0.669432363 | FALSE |
| 103 | 0.520711445 | 0.516780338 | 155299 | 0.478687289 | 0.6952363 | FALSE |
| 26 | 0.428742237 | 0.431501993 | 147799 | 0.477865843 | 0.6952363 | FALSE |
| 88 | 0.51734497 | 0.515069564 | 155123 | 0.499592116 | 0.711111846 | FALSE |
| 118 | 0.556706639 | 0.555528245 | 155109 | 0.501275564 | 0.711111846 | FALSE |
| 97 | 0.396869508 | 0.395688578 | 154889 | 0.528119422 | 0.740581258 | FALSE |
| 104 | 0.495131265 | 0.491622284 | 154700 | 0.551751848 | 0.747930283 | FALSE |
| 105 | 0.527692003 | 0.52411098 | 154753 | 0.545072665 | 0.747930283 | FALSE |
| 73 | 0.570899536 | 0.569681593 | 154745 | 0.546078266 | 0.747930283 | FALSE |
| 112 | 0.440959294 | 0.44430982 | 148498 | 0.563568235 | 0.755553018 | FALSE |
| 55 | 0.520397063 | 0.519155655 | 154412 | 0.588733218 | 0.780711441 | FALSE |
| 71 | 0.558943098 | 0.559026898 | 154323 | 0.600388676 | 0.782132674 | FALSE |
| 58 | 0.5015899 | 0.500024621 | 154306 | 0.602626814 | 0.782132674 | FALSE |
| 29 | 0.536717511 | 0.535019324 | 154154 | 0.622803016 | 0.799810189 | FALSE |
| 4 | 0.161767856 | 0.160670375 | 154062 | 0.635155309 | 0.807176539 | FALSE |
| 34 | 0.465648762 | 0.463176641 | 153898 | 0.657426652 | 0.826866511 | FALSE |
| 33 | 0.448458013 | 0.447102458 | 153690 | 0.686114597 | 0.845514958 | FALSE |

| | | | | | | |
|---|---|---|---|---|---|---|
| 94 | 0.415298597 | 0.413454625 | 153722 | 0.681670215 | 0.845514958 | FALSE |
| 92 | 0.431356641 | 0.432179763 | 149594 | 0.711172162 | 0.847370962 | FALSE |
| 27 | 0.360663186 | 0.362522592 | 149486 | 0.696014231 | 0.847370962 | FALSE |
| 83 | 0.540463718 | 0.542152534 | 149614 | 0.713991977 | 0.847370962 | FALSE |
| 113 | 0.46227646 | 0.463068308 | 149624 | 0.715403353 | 0.847370962 | FALSE |
| 37 | 0.49850252 | 0.49986984 | 149728 | 0.73013856 | 0.856508695 | FALSE |
| 24 | 0.539643123 | 0.539501373 | 149810 | 0.741827676 | 0.861933109 | FALSE |
| 60 | 0.401030939 | 0.402363572 | 149945 | 0.761201178 | 0.876099469 | FALSE |
| 117 | 0.55480554 | 0.556282439 | 150079 | 0.780580418 | 0.890007579 | FALSE |
| 95 | 0.546489355 | 0.546505101 | 150186 | 0.796154017 | 0.899359167 | FALSE |
| 28 | 0.380228306 | 0.381367202 | 150434 | 0.832553475 | 0.931848843 | FALSE |
| 47 | 0.480804161 | 0.480649501 | 152404 | 0.872144794 | 0.962875281 | FALSE |
| 56 | 0.530969439 | 0.530652269 | 150740 | 0.877957183 | 0.962875281 | FALSE |
| 59 | 0.403839622 | 0.404265144 | 150819 | 0.889750856 | 0.962875281 | FALSE |
| 116 | 0.561585701 | 0.559972603 | 152272 | 0.891843498 | 0.962875281 | FALSE |
| 96 | 0.545317615 | 0.544855094 | 150943 | 0.908310758 | 0.972051863 | FALSE |
| 89 | 0.370266596 | 0.369732521 | 152093 | 0.918660826 | 0.974579311 | FALSE |
| 36 | 0.427992568 | 0.427258317 | 151914 | 0.94557101 | 0.985980027 | FALSE |
| 50 | 0.551582116 | 0.55152859 | 151944 | 0.941055747 | 0.985980027 | FALSE |
| 84 | 0.531262675 | 0.530601172 | 151818 | 0.960030893 | 0.987632692 | FALSE |
| 38 | 0.504256642 | 0.503701932 | 151796 | 0.963346643 | 0.987632692 | FALSE |
| 54 | 0.538384669 | 0.538429925 | 151581 | 0.995775101 | 0.996831319 | FALSE |
| 63 | 0.539817242 | 0.53993937 | 151453 | 0.985062767 | 0.996831319 | FALSE |
| 75 | 0.336889097 | 0.335374406 | 151574 | 0.996831319 | 0.996831319 | FALSE |

### METHODS

#### Age balancing

In the entire cohort, a significant age difference between groups was identified (Mann-Whitney U test, $p < 0.001$ ), which would introduce a spurious correlation between age and diagnosis, risking inflating classification performance as subsequent models would exploit age-related patterns rather than diagnosis effects. To mitigate this bias, an age-balancing procedure was applied through data downsampling, both in the sensitivity analysis and in the LOSO-CV within each fold iteration. Specifically, a random subset of subjects, stratifying for site, was removed from the dataset until the statistical test no longer indicated a significant difference, for a critical level of $p>0.1$, achieving a balanced age distribution between the groups. Parallelly, no differences in sex distributions between BD and HC groups were verified (Chi-squared test, $p>0.1$). Thus, all analyses presented in this study were performed with an age-balanced dataset, applied within the specific evaluation framework, which varied across experiments.

#### MRI acquisition and preprocessing

Clinical and structural MRI (sMRI) data was acquired independently in each site, Table S8. The clinical assessment was performed by professional psychiatrists, and diagnosis was based on the Diagnostic and Statistical Manual of Mental Disorders (DSM-IV) criteria for Axis I disorders (SCID-I) *(6)*, with the exception of participants recruited in the Vancouver site (site id=7), for which diagnosis was based on the Mini International Neuropsychiatry Interview *(7)*. The participants' exclusion criteria were: comorbidities, intellectual disability, pregnancy, history of epilepsy, major medical and neurological disorders, neuroleptic treatment in the last 3 months, drug or alcohol abuse in the last 6 months, and medical conditions affecting the immune system.
*sMRI data* was preprocessed using SPM12 (version 7771) *(8)* available at *http://www.fil.ion.ucl.ac.uk/spm/software/spm12/*) and CAT12 version 12.7 *(9)* software in Matlab R2018a (The Mathworks, Inc.®) environment. Firstly, T1-weighted images underwent a visual quality check and were converted from DICOM to NIFTI format. We used the CAT12 pre-processing modules included in the standard voxel-based morphometry (VBM), including affine registration to Montreal Neurological Institute (MNI) space, followed by high-dimensional diffeomorphic normalization using geodesic shooting. Resulting images were fed to brain tissue segmentation, bias correction of intensity non-homogeneities, spatial normalization, modulation with the Jacobian determinant derived from the previous step to preserve the total amount of signal from each region, and spatial smoothing with a 3D Gaussian kernel with a full-width half maximum of 6 mm. Each subject's total intracranial volume (TIV) was estimated. The preprocessed grey matter images were parcelled with the neuromorphometrics brain atlas *(10)* into 122 ROIs, and GMV was corrected for TIV by normalization, which aimed at removing the biasing effect of sex encoded in the GMVs.

**Table S8.** StratiBip network information and MRI acquisition details.

| ID | Reference PI | Scanner | Sequence | Matrix Size | Voxel Size ($m^3$) |
|---|---|---|---|---|---|
| **1-AUOV-Verona Italy** | Marcella Bellani and Paolo Brambilla | Magnetom Allegra Syngo (Siemens, Erlangen, Germany) | T1-MPRAGE | 256x256x160 | 1.00x1.00.x1.00 |
| **2-FSL_ROME - Fondazione IRCCS Santa Lucia, Roma, Italy** | Fabrizio Piras | Philips Achieva 3T (Philips, Best, the Netherlands) | T13D-MPRAGE | 432x432x190 | 0.542x 0.524.x 0.900 |
| **3-JUH-University of Jena, Germany** | Igor Nenadic | Siemens Tim Trio (Siemens, | T1 Magnetization Prepared Rapid | 256x256x192 | 1.00x1.00.x1.00 |

| | | | | | |
|---|---|---|---|---|---|
| | | Erlangen, Germany) | Gradient Echo (MP-RAGE) | | |
| **4-MI-Milano Policlinico, Italy** | Paolo Brambill a | Philips Achieva 3T (Philips, Best, the Netherland s) | T1-Turbo Field Echo (TFE) 3D | 240x240x16 5 | 1.1x1.05x1.05 |
| **5-OSR-Ospedale San Raffaele, Milan, Italy** | Francesc o Benedetti | Philips Intera (Philips, Best, the Netherland s) | T1-Fast Field Echo (FFE) | 256x256x22 0 | 0.9x0.9x0.8 |
| **6- PITTS-Pittsburgh, US** | Mary Philips | 3T Siemens Tim Trio | - | 192x256x19 2 | 1.00x1.00x1.0 0 |
| **7-UBC-University of British Columbia, Vancouver, Canada** | Lakshmi Yatham | Philips Achieva (Philips, Best, the Netherland s) | 3D TFE | 256x256x18 0 | 1.00x1.00x1.0 0 |

**Graph Neural Networks**

The principle underlying GNNs is the message-passing algorithm, which consists of several key steps as shown in Fig. S11. Firstly, in the aggregation step, for each node $\boldsymbol{n_i}$, the node feature information contained in neighbouring nodes within a $\boldsymbol{k}$-hop distance is aggregated and used to generate a message that is passed to the current node $\boldsymbol{n_i}$. Secondly, this message is combined with the current node embedding $\boldsymbol{x_i}$, generating an updated embedding. At this point, a non-linearity, such as ReLU or sigmoid, is applied to the embedding, generating the new embedding $\boldsymbol{h_i^k}$. This process is performed iteratively in each GNN layer $\boldsymbol{L}$, allowing nodes to accumulate information from an increasingly larger neighbourhood, as $\boldsymbol{L = k}$. Finally, in tasks like graph classification, where the goal is to obtain a single representation for the entire graph, a readout function is applied after the last GNN layer to aggregate node embeddings into a graph-level embedding.
In this study we compared three types of GNN architecures: graph convolutional networks (GCN), graph attention networks, and graph isomorphism networks.

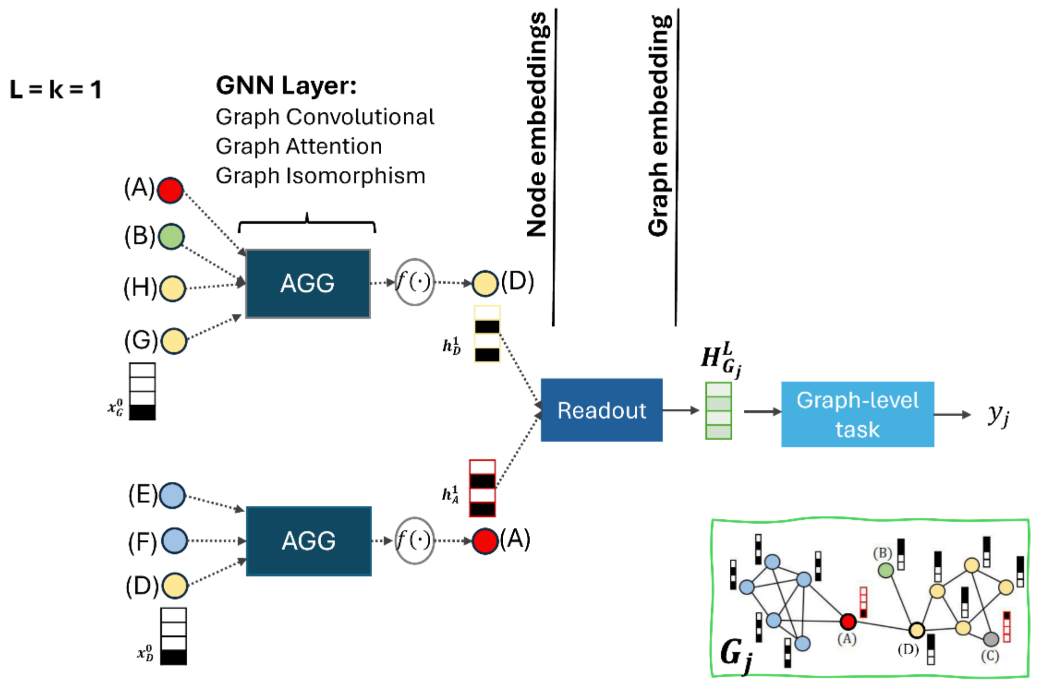

**Fig. S11.** GNNs' message-passing and architecture scheme. Neighboring nodes of A and D are selected and their messages, $\boldsymbol{x_n^0}$, are transformed and aggregated before updating the current node embedding, $\boldsymbol{h_n^1}$. A readout layer is used to transform all the node embeddings into a graph embedding, $\boldsymbol{H_G^L}$, and a prediction is outputted, $\boldsymbol{y_j}$. L: number of layers; k= hop-distance from current node to neighboors; j: current subject; , $\boldsymbol{G_j}$: graph of current subject ;AGG: aggregation function; $\boldsymbol{f}(\cdot)$: non-linear function.

**Graph Convolutional Network.** Graph convolutional networks (GCN) were introduced by Kipf and Welling in 2017 *(11)*, initially intended for semi-supervised node classification but later extended to graph classification. In this case, the message transformation is linear, given by message computation $m_{u\to v} = Wx_v^L$ , where $W$ is the matrix of learnable weights, and $u$ is a neighbouring node of $v$. The message is then normalized by the degree of the nodes involved and aggregated across all k-hop neighbouring nodes. Thus, in a GCN, the neighbour and message importance are determined by the graph topology as the message is weighted by the node's degree.

**Graph Attention Network.** Graph attention networks (GATs) *(1)* introduced attention mechanisms in GNNs. In this case, learnable attention scores dynamically determine the importance of each neighbour $u$ to a node $v$, defined as $\alpha_{u\to v} = f([m_u | m_v])$, where $m_u$ and $m_v$ are the messages of the respective nodes, which are concatenated and transformed by the attention mechanism $f(.)$, usually a *LeakyReLU* activation followed by a node-wise *softmax* normalization. Then, the neighbour messages aggregation is weighted according to the attention score, $\alpha_{u\to v}$.

**Graph Isomorphism Network.** Graph isomorphism networks (GIN) *(2)* were designed to be as powerful as the Weisfeiler-Lehman (WL) graph isomorphism test. This was done by passing the aggregated neighbourhood information, $agg = \sum_i^N m_{i\to v}$ , where $m_{i\to v}$ is the message passed from neighbour node $i$ to $v$, through a multi-layer perceptron, $MLP(agg)$. The latter makes the overall message update function injective, ensuring different neighbourhood structures produce different node embedding representations and increasing the expressiveness of the model.

#### Model Validation Framework: LOSO-CV

In the LOSO-CV, iteratively, in each fold, data from one site were held out as a test set while all others were used as a training set. First, we performed hyperparameter tuning and model selection in a 5-fold CV with the training set, and then the best model was re-trained and tested on data from the hold-out site. Within the 5-fold CV, the training set was split into 5 different folds, each with a training and validation portion, denoted as an internal training and validation set. The GNN model was trained in the internal training set and evaluated in the internal validation set with varying model configurations. By comparing all the results on the internal validation set, the best model configuration is chosen and moved to the outer fold, the test phase. In the test stage, the best model is retrained in the entire training set and tested in the hold-out data. All sites, except site 2, were used as a hold-out test set once. The age balancing procedure was applied within each fold to the training set as well as the multi-site data harmonization, which was applied both at the 5 internal folds of the CV, as well as in the outer fold of the LOSO-CV, to control for site-effects in training, validation, and test sets.

#### Multi-site ComBat Harmonization

MIND networks were directly harmonized with ComBat model *(12)*, which has been previously validated for harmonizing several neuroimaging meta-features, such as brain functional connectivity matrices *(13)*, and structural features like volumes and cortical thickness *(14, 15)*. ComBat is a statistical method that uses an empirical Bayes framework to correct batch effects. It adjusts data by removing variation associated with non-biological factors such as site, while preserving variability related to variables of interest, such as biological covariates, age, sex, and diagnosis *(15)*. Briefly, for a given feature $v$ , subject $j$ and site $i$, the observed data $y_{ijv}$ is modelled as:

$$\boldsymbol{y_{ijv} = \alpha_v + X_{ij}^T\beta_v + \gamma_{iv} + \delta_{iv}\epsilon_{ijv}} \quad (1)$$

Where $\alpha_v$ is the standardisation coefficient of feature $v$ across all subjects and sites, $X_{ij}^T$ the biological covariate vector, and $\beta_v$ the feature-specific regression coefficients, $\gamma_{iv}$ and $\delta_{iv}$ the additive and multiplicative site effects, respectively, and $\epsilon_{ijv}$ the error term. Then, the harmonised features are computed by removing the estimated site effects:

$$\boldsymbol{\hat{y}_{ijv}^{ComBat} = \frac{\hat{\sigma}_v}{\hat{\delta}_{iv}}(y_{ijv}^{stand} - \hat{\gamma}_{iv}) + \hat{\alpha}_v + X_{ij}\hat{\beta}_v} \quad (2)$$

Where the standardised data is represented as:

$$y_{ijv}^{stand} = \frac{y_{ijv} - \hat{\alpha}_v - X_{ij}^T \hat{\beta}_v}{\hat{\sigma}_v} \quad (3)$$

For more details, please refer to the original paper *(12)*. The ComBat model was applied within the CV frameworks, respecting the independence between training and test set, with the Python function available in https://github.com/inesws/neurocombat_pyClasse.git.

**In sensitivity analysis.** In this stage, the dataset was split into training, validation, and test sets, stratifying for site, i.e., the proportion of subjects for each site was kept in the three subsets. ComBat was then applied as in *(16)*, where site parameters were exclusively estimated in the training set and then applied to the whole dataset, including validation and test set, effectively avoiding data leakage.

**In LOSO-CV.** For each of the CV folds, modified (M-)ComBat *(4)* was used to harmonize data from an unseen independent site. First, in each fold iteration, the training set, composed of all sites except one, was harmonized with ComBat. Then, after model training, the hold-out site used for testing was harmonized with the harmonized training set using M-ComBat *(5)*. In this variant of ComBat, the model estimates the site differences between a reference site and a new site, and shifts all sites to the mean and variance of the reference site. In this study, the harmonized training set served as a reference, as we assumed it as a homogeneous site, and it remains unchanged, while the hold-out test set is shifted to the mean and variance of the harmonized training set during the model testing phase. The latter efficiently avoids any data leakage in the training set and allows for homogenizing training with test sets. In this case, the new standardization is performed site-wise, and the correction given by:

$$\hat{y}_{ijv}^{ComBat} = \frac{\hat{\sigma}_{REF,v}}{\hat{\delta}_{iv}} (y_{ijv}^{stand} - \hat{\gamma}_{iv}) + \hat{\alpha}_{REF,v} + X_{ij} \hat{\beta}_v \quad (4)$$

Where $i = reference\ site$ in $\alpha_{REF,v}$ and $\hat{\sigma}_{REF,v}$. For more details, please consult the original paper *(4)*.
